# Genome-wide Analysis Identifies Novel Gallstone-susceptibility Loci Including Genes Regulating Gastrointestinal Motility

**DOI:** 10.1101/2021.07.16.21260637

**Authors:** Cameron J Fairfield, Thomas M Drake, Riinu Pius, Andrew D Bretherick, Archie Campbell, David W Clark, Jonathan A Fallowfield, Caroline Hayward, Neil C Henderson, Andrii Iakovliev, Peter K Joshi, Nicholas L Mills, David J Porteous, Prakash Ramachandran, Robert K Semple, Catherine A Shaw, Cathie LM Sudlow, Paul RHJ Timmers, James F Wilson, Stephen J Wigmore, Athina Spiliopoulou, Ewen M Harrison

## Abstract

**Objective:** Genome-wide association studies (GWAS) have identified several risk loci for gallstone disease. As with most polygenic traits, it is likely many genetic determinants are undiscovered. The aim of this study was to identify novel genetic variants that represent new targets for gallstone research and treatment.

**Design:** We performed a GWAS of 28,627 gallstone cases and 348,373 controls in the UK Biobank and a GWA meta-analysis (43,639 cases and 506,798 controls) with the FinnGen cohort. We assessed pathway enrichment using gene-based then gene-set analysis and tissue expression of implicated genes in Genotype-Tissue Expression project data. We constructed a polygenic risk score (PRS) and evaluated phenotypic traits associated with the score.

**Results:** Seventy-five risk loci were identified (P<5*10^−8^) of which forty-six were novel. Pathway enrichment revealed associations with lipid homeostasis, glucuronidation, phospholipid metabolism and gastrointestinal motility. *ANO1* and *TMEM147*, both in novel loci, are strongly expressed in the gallbladder and gastrointestinal tract. Both regulate gastrointestinal motility. The gallstone risk allele rs7599-A leads to suppression of hepatic *TMEM147* expression suggesting the protein protects against gallstone formation. Individuals in the highest decile of the PRS demonstrated a 6-fold increased risk of gallstones compared to the lowest risk category. The PRS was strongly associated with increased body mass index, serum liver enzyme and C-reactive protein concentrations and decreased lipoprotein cholesterol concentrations.

**Conclusion:** This GWAS demonstrates the polygenic nature of gallstone risk and identifies 46 novel susceptibility loci. For the first time, we implicate genes influencing gastrointestinal motility in the pathogenesis of gallstones.

**Summary Box:** *What is already known on this subject?:*

- Genome-wide association studies (GWAS) have identified 29 genetic variants within independent loci which increase the risk of gallstone disease.
- Most of these variants lie within or near to genes that regulate lipid or bile acid metabolism. Gallstones are known to have a significant genetic component with 25-50% of gallstone disease due to genetic risk factors.
- Much of this risk is not accounted for by the known gallstone-susceptibility loci.

*What are the new findings?:*

- We performed a GWAS in the UK Biobank (28,627 gallstone cases, 348,373 controls) and a GWA meta-analysis (43,639 cases and 506,798 controls) with the FinnGen cohort.
- We identified a total of 75 gallstone-susceptibility loci with 46 of these being new and the remaining 29 being those already identified.
- We annotated the variants based on their position within or near to genes and assessed pathway enrichment through gene-set analysis.
- We identify two novel gallstone-susceptibility loci in which the lead variants lie within genes governing gastrointestinal motility which highly expressed in gallbladder (*ANO1* and *TMEM147*).
- We demonstrate further loci involved in primary cilia function.
- We report significant association of a polygenic risk score with gallstones using independent subsets of the study population.

*How might it impact on clinical practice in the foreseeable future?:*

- The genes and pathways identified represent novel targets for development of medication targeting primary or secondary prevention of gallstones. This may be of particular benefit to those unable to undergo cholecystectomy.
- The individual variants or polygenic risk score identified in this GWAS could form the basis for identification of individuals with high risk of gallstones to support screening or treatment of gallstone disease. This screening may be of particular benefit in populations with elevated risk of gallstones such as haemolytic disease or bariatric surgery.

## Introduction

Gallstone disease is one of the most common reasons for acute presentation to hospitals worldwide^1^ and generates the greatest healthcare expenditure of any gastrointestinal condition in the United States^2^. Prevalence of gallstones reaches 20-40% in high-risk populations^3,4^ and gallstones are highly heritable^5^. Identification of novel pathways influencing gallstone development may improve risk stratification in high risk populations and support development of medications targeting gallstone formation and other cholestatic disease for use in primary and secondary prevention.

A total of seven genome-wide association studies (GWAS) have been published revealing 29 gallstone-promoting loci^6–12^. Most gallstones are composed of cholesterol which crystallises in bile at high concentration and/or in bile lacking protective bile acids^13^. Many of the identified variants relate to cholesterol and bile acid metabolism. Despite evidence of impaired gallbladder and gastrointestinal motility contributing to the formation of gallstones, there have been no gallstone-susceptibility variants identified that influence these characteristics^14^. Using larger and more richly phenotyped cohorts may identify such variants and contribute to the knowledge of gallstone formation.

The most recent GWAS and GWAS meta-analysis used publicly-available summary GWAS data from the Global Biobank Engine (GBE)^15^ based on UK Biobank (UKB) data^16^. GBE is based on three-level ICD-10 and ICD-9 codes and considered gallstones to be code “K80” (“cholelithiasis”) or self-reported gallstones at study enrolment. The UKB captures several other informative variables including OPCS codes, other ICD-9 and 10 codes, self-reported procedures relating to gallstones and primary care data. By incorporating these additional data, a greater proportion of patients with gallstone disease can be classified as cases rather than misclassified as controls.

The aim of this study was to perform an updated GWAS using more richly phenotyped participants to identify novel pathways contributing to gallstone formation. The genetic associations identified were replicated in the Generation Scotland: Scottish Family Health Study (GS-SFHS) cohort^17^ and meta-analysed with FinnGen summary association statistics^18^. A polygenic risk score was derived and analysis for gene-environment interaction undertaken.

## Materials and Methods

Methods for identification of participants, determination of gallstone status and genotyping for the UKB, GS-SFHS and the FinnGen cohort are provided in Supplementary Materials 1. Diagnostic codes used for the identification of gallstones are shown in Supplementary Tables 1-3. The UKB received ethical approval (Research Ethics Committee [REC] reference number: 11/NW/0382). UKB data access was approved under projects 30439 (phenotype data) and 19655 (genotype data). Ethical approval for GS-SFHS was granted by NHS Tayside REC (reference number: 05/S1401/89). The FinnGen study protocol was approved by the Ethical Review Board of the Hospital District of Helsinki and Uusimaa (Nr HUS/990/2017).

### Genome-wide Association Study

#### Initial Analysis

Genotyped and imputed SNPs were analysed. Participants of self-reported European ancestry were included and outliers for heterozygosity and unexpected runs of homozygosity were excluded. One randomly-selected participant from each pair of related individuals in the UKB (Kinship > 0.0884^19^) was excluded.

Association with gallstones was analysed using logistic regression adjusted for age, sex, the first 20 genetic principal components and batch, with batch included as a random effect. Imputation dosage was used for imputed SNPs.

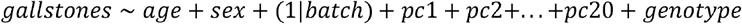

SNPs were excluded based on minor allele frequency (MAF < 0.001), imputation quality (INFO < 0.3) and departure from Hardy-Weinberg equilibrium (HWE: P < 5*10^−6^). The MAF was selected on the basis of ensuring 25 or more minor alleles in the cases (*MAF* = 25/(2 * *n_cases)*) which corresponded to a MAF of 0.0004. This was rounded up to 0.001.

Genome-wide significance was determined as P<5*10^−8^ in the UKB and Bonferroni-corrected *P*_*C*_ = 0.05 in the replication cohorts (0.05 divided by the number of loci tested). Replication was undertaken by analysing the lead SNP within each locus or a linkage disequilibrium (LD) proxy.

P values smaller than 4.94 * 10^−324^ (the limit of numerical precision on a Linux 64-bit operating system) were rounded to 4.94 * 10^−324^ (Supplementary Materials 1).

#### Genome-Wide Association Meta-analysis

A genome-wide association meta-analysis was undertaken using UKB data and FinnGen data in an inverse variance-weighted fixed effects model. SNPs shared between the datasets (10,044,759) were analysed. RsID was converted to the format “rs1234:A:T” to avoid ambiguity at multi-allelic SNPs.

#### Linkage Disequilibrium Clumping and Conditional Analyses

LD clumping was performed using the Functional Mapping and Annotation of Genome-Wide Association Studies (FUMA) web application version 1.3.5d^20^. Loci were established for each lead SNP with a minimum distance of 250Kb between loci and using an *r*^2^ < 0.25 to indicate separate independent SNPs within the same locus. Details of the FUMA parameters are available in Supplementary Materials 2A. Five loci with rare lead SNPs not present in the 1000 Genomes Phase 3 reference panel (MAF_UKB_⍰0.01-0.001) were generated manually by creating a 500 KB window centred on the lead SNP.

Each locus was re-analysed whilst conditioning on the lead SNP and further signals with genome-wide significance were identified. This process was repeated with additional lead SNPs until no remaining SNPs reached genome-wide significance.

#### Population Stratification

The genomic inflation factor *λ*_*gc*_ was calculated using the summary GWAS statistics. Evidence of test statistic inflation in *λ*_*gc*_ was investigated with linkage disequilibrium score regression^21^.

### Sensitivity Analysis

As the definition of gallstones was based on clinical detection of gallstones it is possible that detection bias may influence the results. Gallstones detected during investigations for other conditions may be recorded despite causing no symptoms. Genetic variants causing other hepatic pathology may therefore influence the likelihood of detection, rather than formation, of gallstones. Individuals requiring surgical management of gallstones are likely to be symptomatic, minimising detection bias. A sensitivity analysis in which cases underwent a cholecystectomy, or any gallstone-specific procedure as defined in Supplementary Table 2, and controls were free from gallstone disease was undertaken at each of the identified lead SNPs. Those with gallstones who did not undergo surgery were excluded.

### Association of Lithogenic Loci with Phenotypic Traits

Associations with serum biochemistry values for each identified variant were assessed in an age- and sex-adjusted linear regression model. Serum bilirubin (total, conjugated and unconjugated), alanine aminotransferase (ALT), aspartate aminotransferase (AST), gamma-glutamyl transferase (GGT), alkaline phosphatase (ALP), total cholesterol (TC), low-density lipoprotein cholesterol (LDL-C), high-density lipoprotein cholesterol (HDL-C), triglycerides, apolipoprotein A and B (ApoA and ApoB), lipoprotein A (LipoA), haemoglobin (Hb), red blood cells (RBC), reticulocyte percentage, C-reactive protein (CRP) and Cystatin C were assessed. Each trait was assessed visually via histogram and log-transformed in the case of skewed distribution.

### Gene-Environment Interaction

The *UGT1A1* polymorphism rs4148325 associates with bilirubin and causes Gilbert’s syndrome but is a comparatively modest driver of gallstones in GWAS^12,22^. In patients with haemolytic disease or cystic fibrosis, the variant is a significant risk factor for gallstones^23–25^. This raises the possibility of a gene-environment interaction in which defects of bilirubin metabolism only lead to gallstone formation in the context of increased bilirubin production or decreased bilirubin excretion. Each identified locus was re-analysed for a gene-environment interaction with serum unconjugated bilirubin as a covariate and as an interaction term with genotype. A Wald test was performed to determine the presence of a gene-environment interaction and significance was determined using a Bonferroni correction. This was not conducted at the known *UGT1A1* locus as unconjugated bilirubin is likely to be a mediator of the relationship between the variant and gallstones.

### Polygenic Risk Score

A polygenic risk score (PRS) was derived using a random 80%/20% split of the UKB cohort. A separate GWAS was conducted in the 80% and the summary associations statistics used to derive the PRS which was validated in the 20% split using PRSice^26^. This was conducted as an additive model with default PRSice parameters on directly genotyped SNPs. The PRS was evaluated for association with the same phenotypic traits as individual risk loci in a simple, unadjusted linear or logistic regression.

### Pathway Enrichment

We conducted a gene-based analysis in MAGMA to identify genes and gene-sets showing significant association with gallstones. Methods for gene-based and gene-set analysis have been described previously^27^. In brief, summary association statistics, following FUMA-based annotation, were grouped by gene and the average χ^2^ statistic across all SNPs in each gene computed and converted to gene-based P values. Genes demonstrating significant association (after Bonferroni correction) were taken forward for a competitive gene-set analysis in which gene-sets were tested for association with the phenotype. This tests whether the mean association of the phenotype with genes in the gene set is greater than that of genes not in the gene set. The mean association was adjusted for gene size, gene density (based on genetic distance using between-SNP LD), the inverse of the mean minor allele frequency in the gene, and the log value of these three variables. This adjustment is necessary to prevent spurious associations driven by the number and frequency of possible mutations within a given gene rather than a true effect. 4,761 curated gene sets and 5,917 gene ontology (GO) terms were available for analysis from Molecular Signatures Database v6.2. Gene tissue expression analysis was undertaken in MAGMA using the identified genes. Loci without clear relation to gallstones were assessed by literature search based on the FUMA gene annotations for genes nearest to the lead SNP to identify further pathways not present or identified within the curated gene-sets. Lead SNPs were analysed in the Genotype-Tissue expression project (GTEx) to assess their effect on expression of putative causal genes.

### Plotting and Statistical Analysis of Results

Genomic analyses were performed using SNPtest (version 2.5.4 for gene-environment interaction model, version 2.5.2 for all other analyses) and QCTOOLS (version 2.0.6)^28^ within a Linux high-performance compute cluster. GWA meta-analysis was undertaken in METAL^29^. Post-GWAS analysis, regression analyses and plotting were performed using R version 3.6.3^30^. Methods and results are reported in accordance with the STREGA guidelines^31^ (Supplementary Materials 3).

### Patient Involvement

During the design stage of the project, the NHS Lothian Public and Patient Involvement and Engagement was involved in drafting lay summaries of the proposed research. Published results will be returned to the UK Biobank team who inform participants of research outputs.

## Results

### Gallstones Cohort

A total of 502,616 participants entered the UKB study of which 377,000 were taken forward for GWAS. A history of gallstones was present in 28,627 with 348,373 controls. The median follow-up available was 9.0 years (range 7.3-11.9). Baseline demographics of the UKB cohort are shown in Supplementary Table 4.

A total of 24,096 participants entered GS-SFHS of which 6,317 unrelated individuals were taken forward for GWAS. A history of gallstones was present in 1,089 with 5,228 controls. The mean follow-up available was 10.7 years (range 9.6-14.7) (Figure 1).

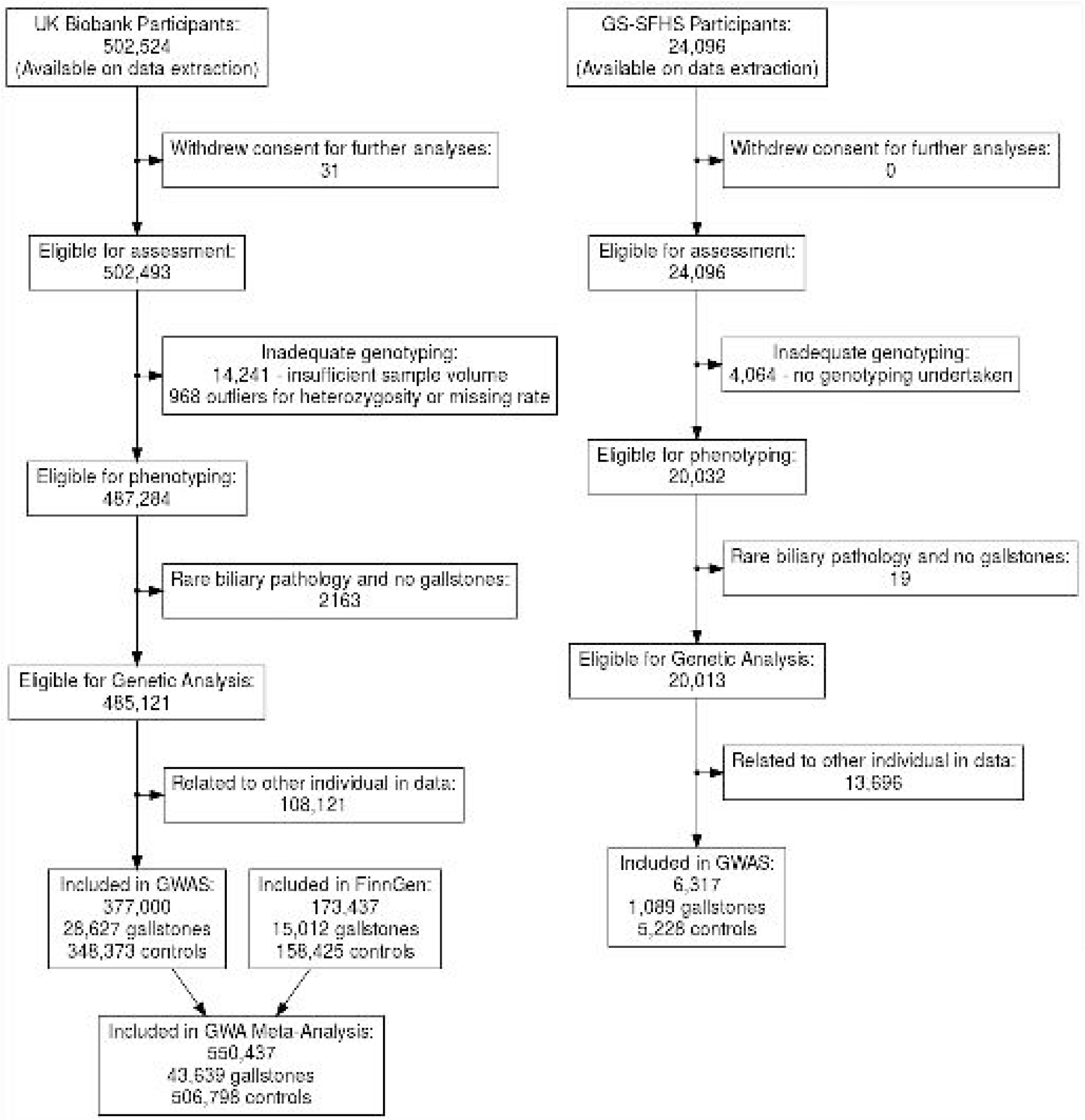

In FinnGen 173,437 participants were analysed for gallstones (15,012 cases, 158,425 controls).

### Genome-wide Association Study

#### Initial UKB Analysis

A total of 4,065 SNPs were identified with genome-wide significant P values after removal of low-quality SNPs (see Supplementary Materials 4). An additional 11,539 SNPs demonstrated borderline significance (*P* < 5 * 10^−5^). LD-clumping revealed 50 loci with at least one significant signal (see Supplementary Materials 2B). Five additional loci contained variants with a significant association with gallstones for which the lead SNP was not present in the 1000 Genomes reference panel (MAF 0.01 to 0.001) and were therefore not annotated by the FUMA application. LD-clumping for those loci was performed using boundaries of 250KB from the lead SNP.

Of the 55 loci identified in the UKB, 27 loci are newly identified variants, 7 are known from previous GWAS using other datasets and 21 variants were previously identified only in studies using GBE data^10,12^ (Table 1). Four variants were replicated in GS-SFHS (rs11887534, rs1993453, rs1260326 and rs62129966; P < 0.00091, corrected for 55 variants). 33 variants from the UKB GWAS reached the replication threshold in the FinnGen cohort (P < 0.00012, corrected for 42 variants available) and 38 reached nominal significance (*P* < 0.05). 10 of the 27 novel loci identified in the UKB GWAS reached the replication threshold in FinnGen. Results for replication in GS-SFHS and FinnGen at each of the 55 loci identified in the UKB are shown in Supplementary Table 5. Loci with replication in either GS-SFHS or FinnGen are highlighted in bold in Table 1.

**Table 1:**
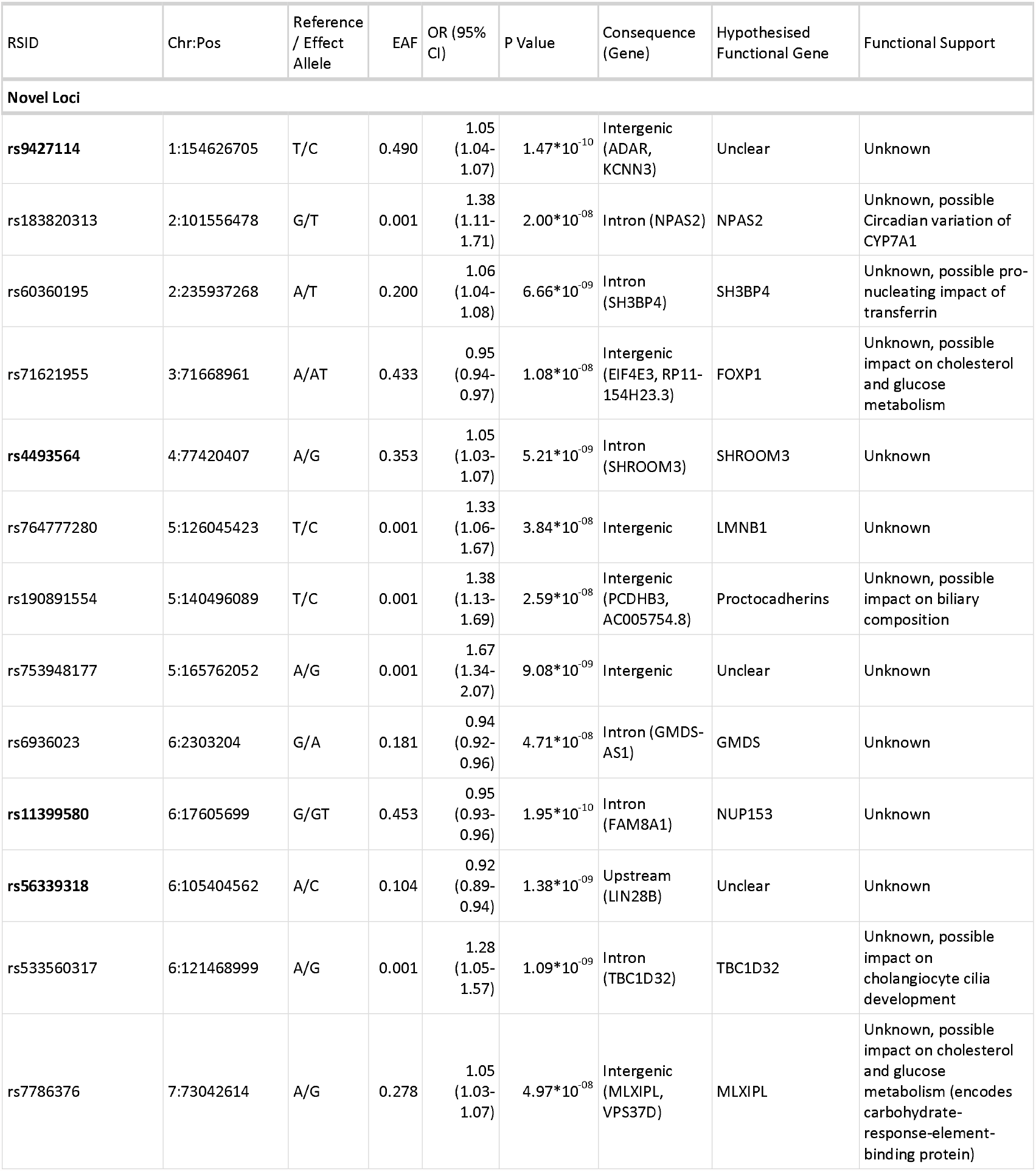
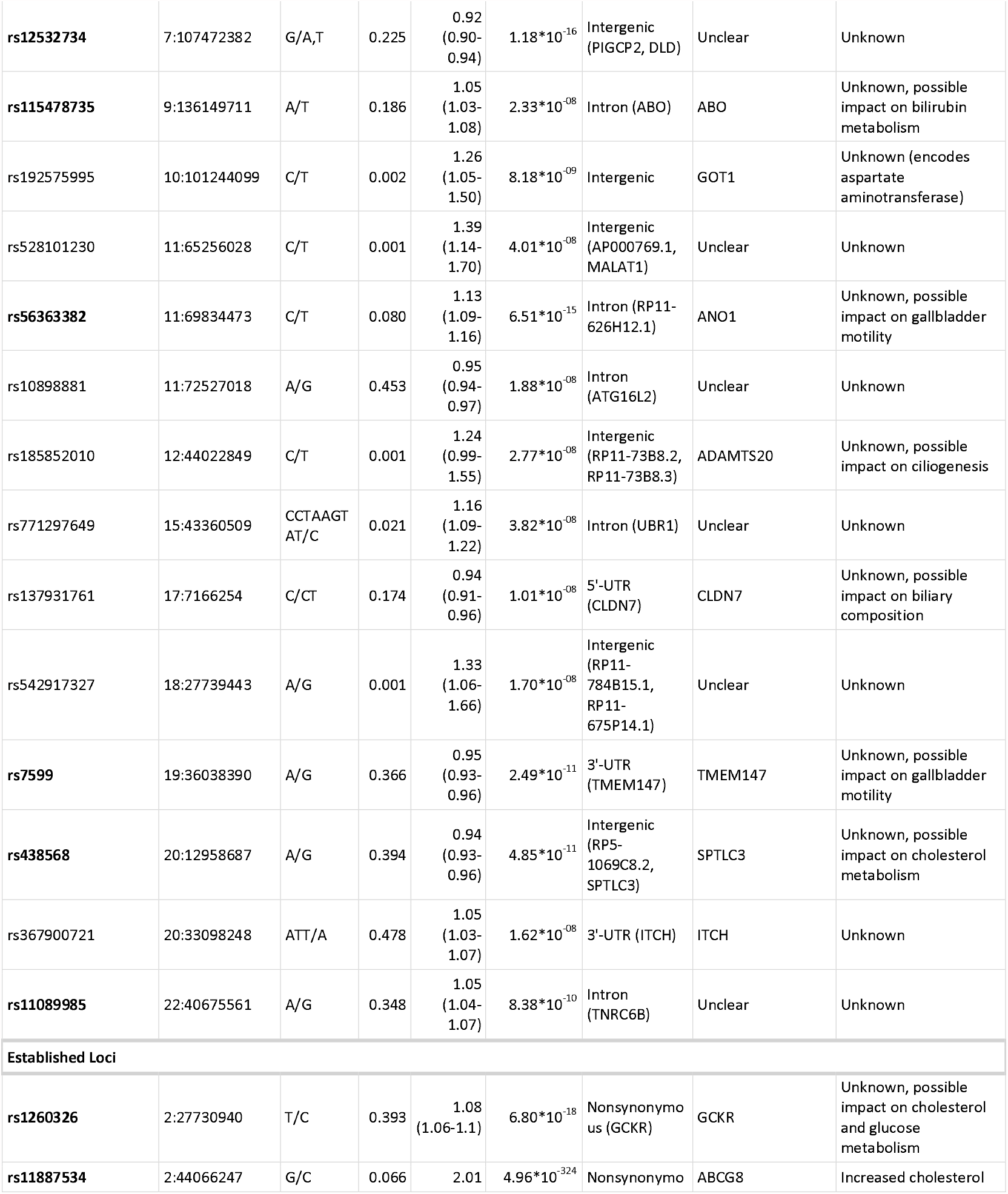
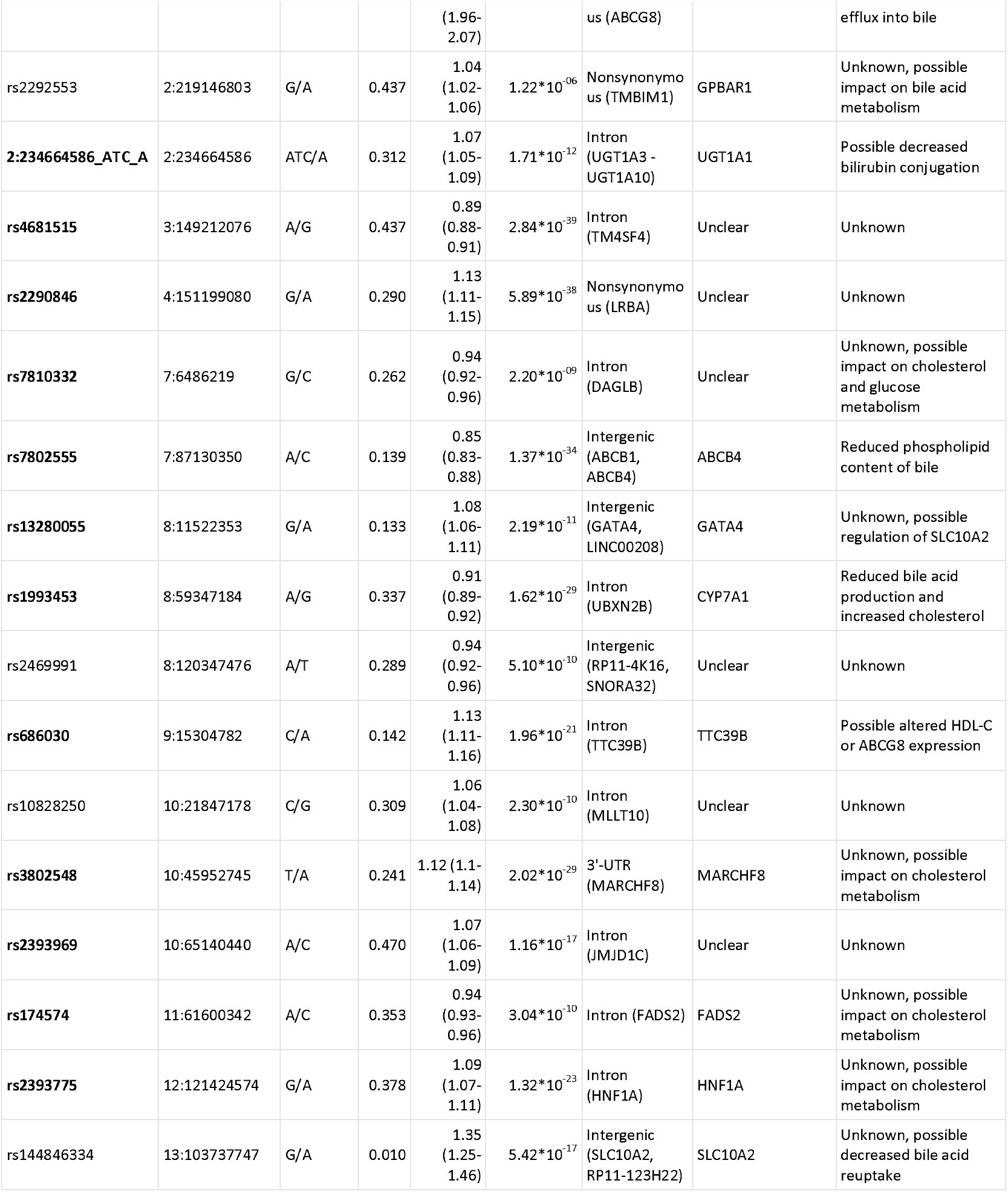
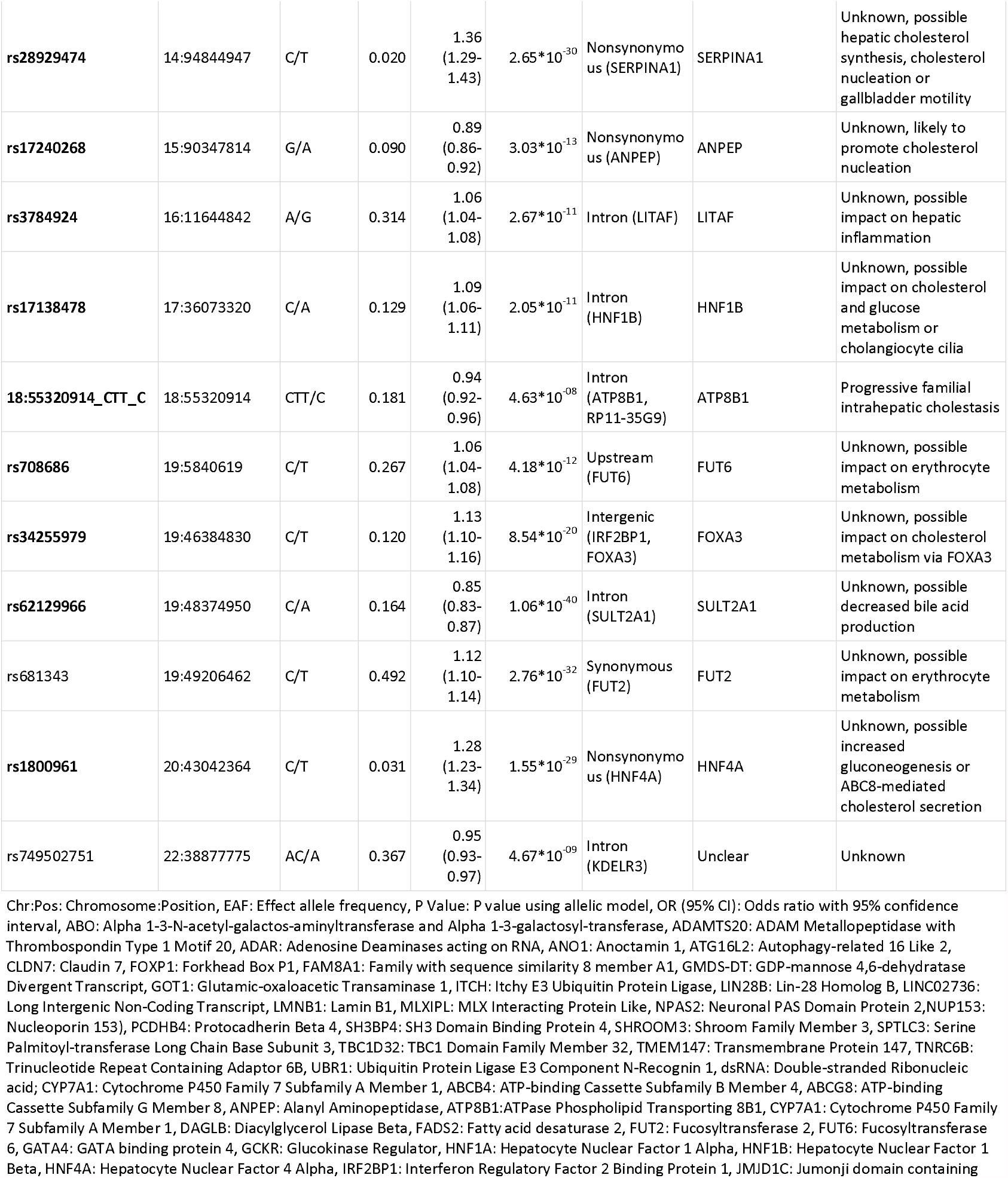
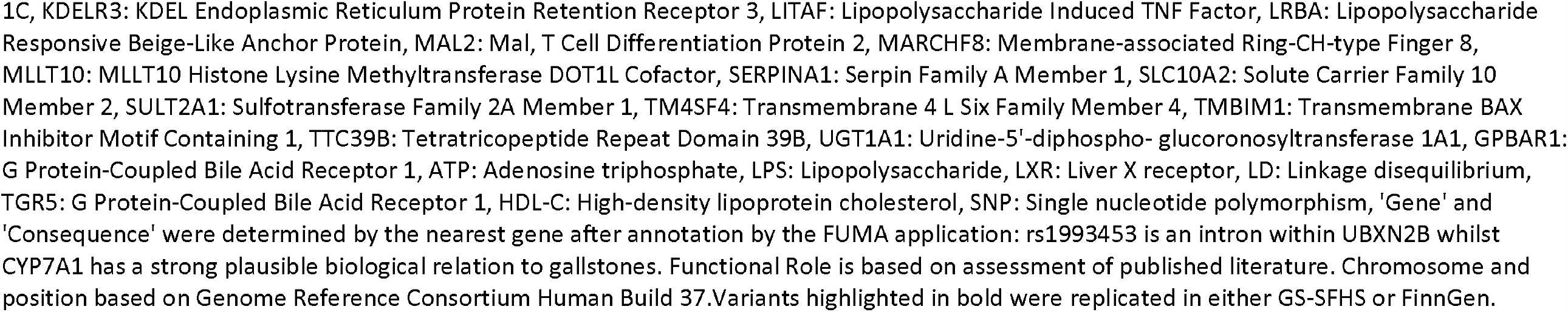
Gallstone-Susceptibility Loci in the UK Biobank GWAS

The D19H variant of *ABCG8* (rs11887534) demonstrated a highly significant result (*P* < 4.94 * 10^−324^). 23 adjacent variants demonstrated identical P values and almost identical *β*-coefficients. As rs11887534 is a known protein-altering variant (D19H) and previously identified as the lead SNP by other GWAS, this was considered as the lead SNP.

#### GWA Meta-analysis

GWA meta-analysis with FinnGen data revealed a total of 63 loci associated with gallstone disease (*P* < 5 * 10^−8^). Of these, 43 were identified in the UKB, one was previously identified with GBE data^10^ and a further 19 were novel loci bringing the total number of novel gallstone-susceptibility loci to 46 and the overall total of gallstone-susceptibility loci to 75. Of the 29 previously reported loci, 28 were identified in the UKB GWAS and all 29 were identified in the meta-analysis. Twelve loci identified in the UKB GWAS did not retain significance on meta-analysis. The 63 lead SNPs from the meta-analysis are shown in Table 2.

**Table 2:**
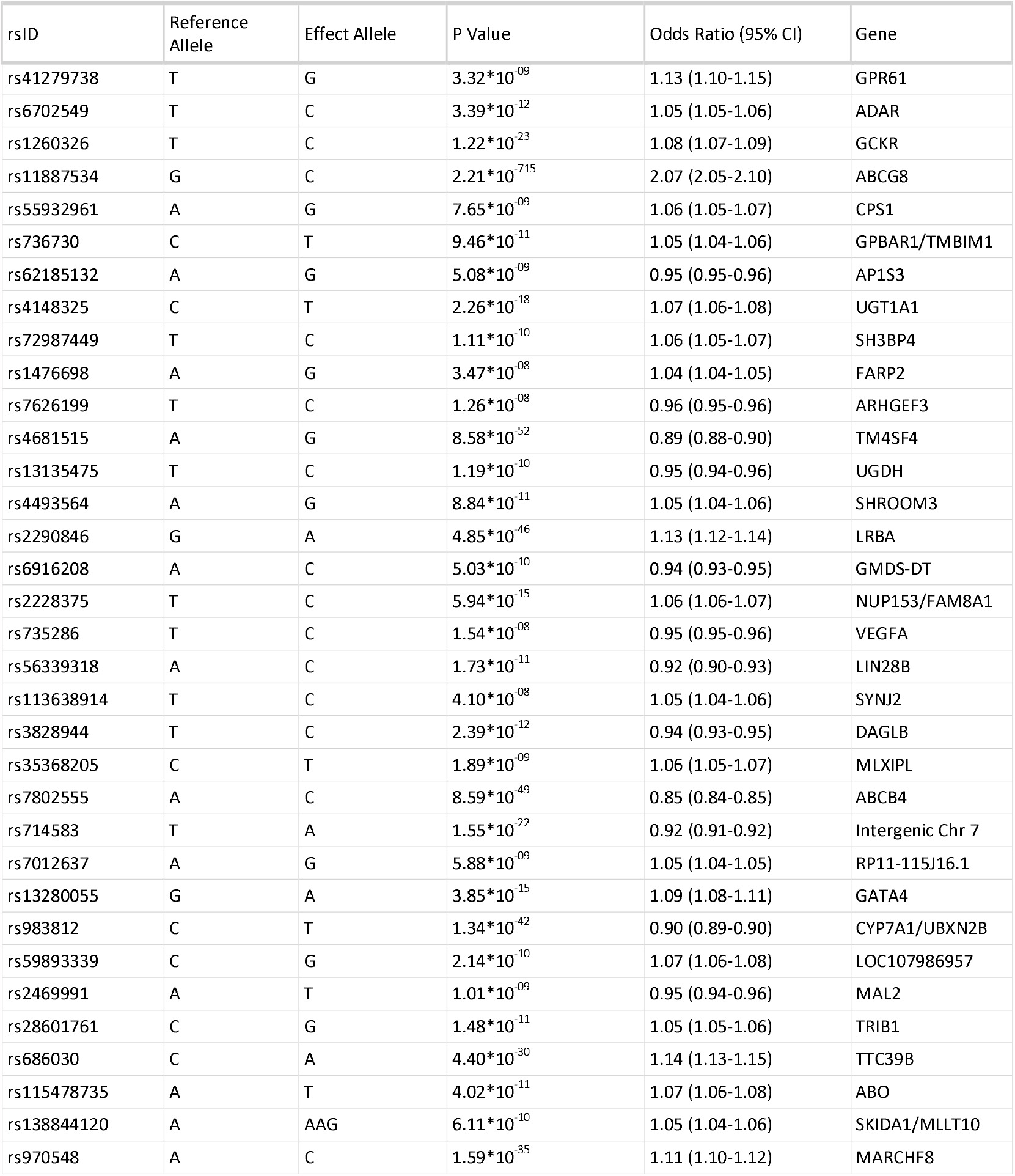
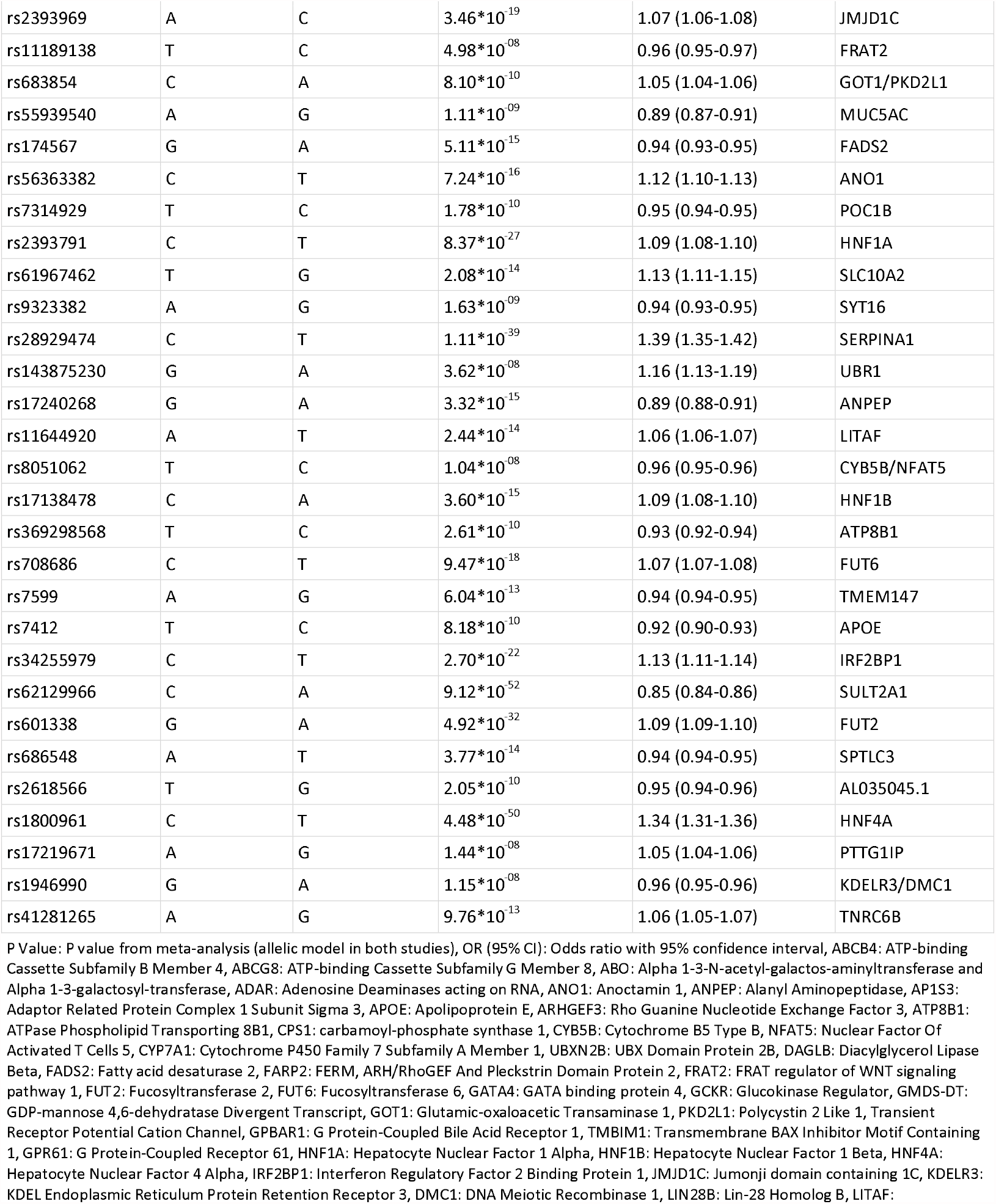
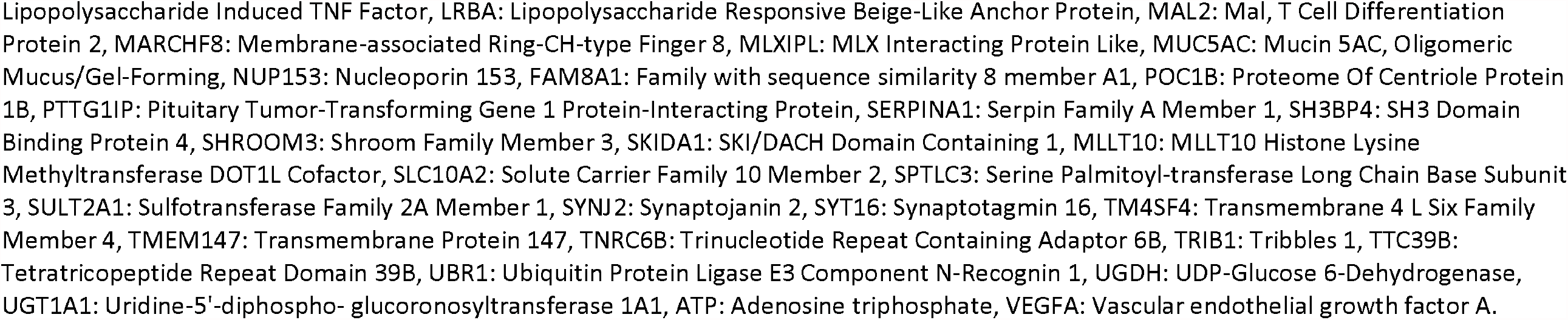
Gallstone-Susceptibility Loci in GWAS Meta-analysis

A combined circular Manhattan plot for association of variants with gallstones in the meta-analysis as well as the 12 loci only identified in the UKB is shown in Figure 2. Rectangular Manhattan plots for the meta-analysis and separately for the initial UKB analysis are available in Supplementary Figures 1 to 4.

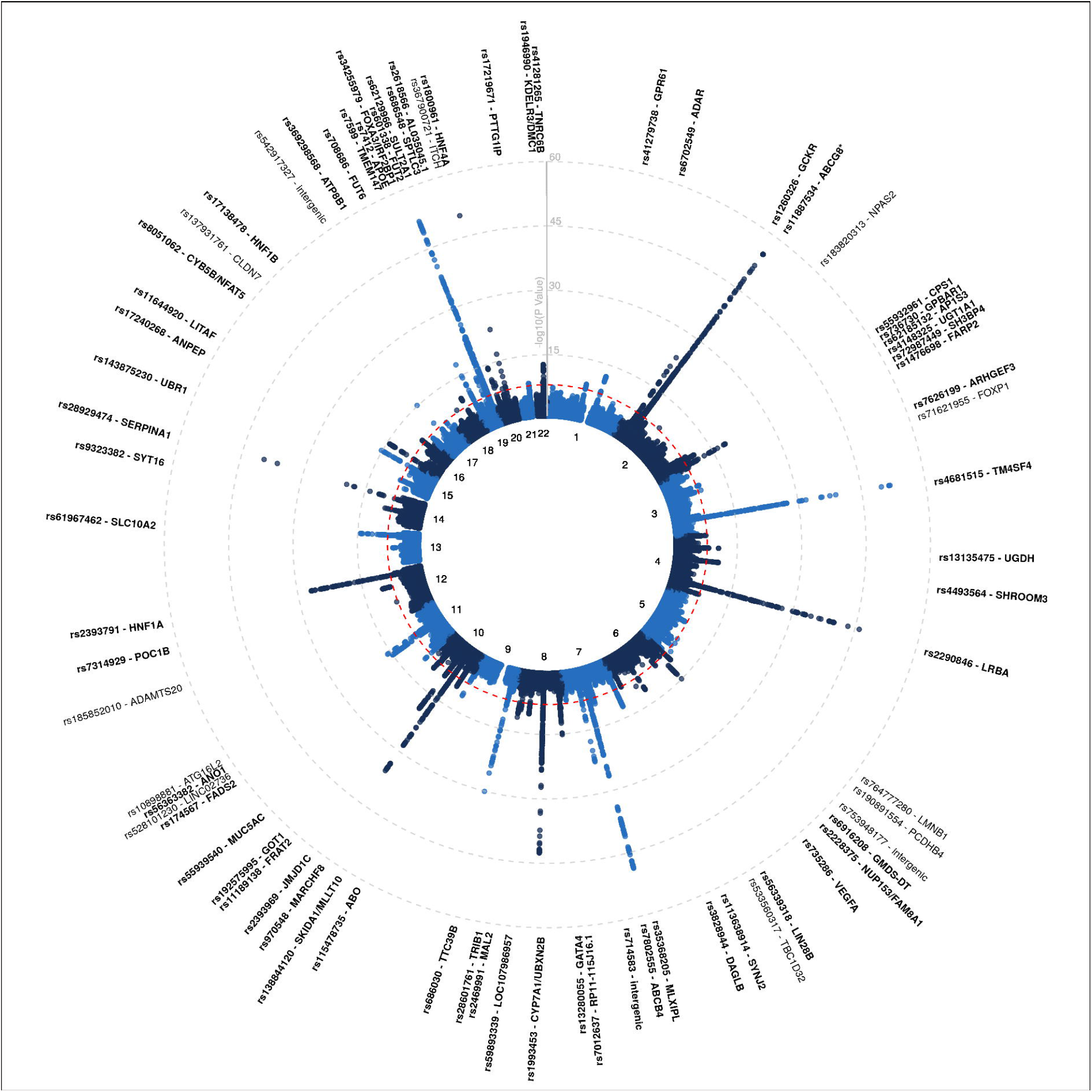

### Conditional Analyses

Conditional analyses, adjusting for the lead SNP, were undertaken for the 55 loci identified in the UKB GWAS. After conditional analyses, a total of 6 loci were found to have additional independent signal suggesting that more than one variant within these loci has a causal relationship with gallstones. The remaining 49 loci did not have any additional SNPs reaching genome-wide significance (*P* < 5 * 10^−8^).

The *ABCG8* locus (rs11887534) contained 10 variants with independent and significant association with gallstones within *ABCG8, ABCG5* and *DYNC2LI1*. The *TM4SF4* locus (rs4681515) contained 2 variants, the *ABCB4* locus (rs7802555) contained 3 variants, the *SLC10A2* locus (rs144846334) contained 3 variants, the *FOXA3* locus (rs34255979) contained 2 variants and the *TNRC6B* locus (rs11089985) contained 2 variants with independent and significant association with gallstones. Full results of the conditional analyses are provided in Supplementary Materials 4. This study did not detect a second independent signal previously identified at the locus containing *TTC39B*^9^ although a more stringent genome-wide significance threshold was used in this study (*P =* 5 * 10^−8^ versus *P* = 6.49 * 10^−4^).

### Population Stratification

After adjusting for the first 20 genetic principal components, there was evidence of test statistic inflation (*λ*_*gc*_ = 1.103). Test statistic inflation is seen in traits caused by multiple genes (polygenicity) but can also be a feature of unmeasured population substructure in which genetically-similar individuals also share similar environmental exposures^32^. Linkage disequilibrium score regression determines the extent to which polygenicity drives the test statistic inflation. The linkage disequilibrium score regression intercept was 1.042 and the proportion of test statistic inflation ascribed to causes other than polygenicity was estimated to be 10.9-17.2% confirming that polygenicity is the main driver of test statistic inflation. *λ*_*gc*_ was similar to the GBE GWAS^12^. The quantile-quantile plot for the UKB is shown in Supplementary Figure 5.

### Sensitivity Analysis

A sensitivity analysis was conducted in each of the 55 lead SNPs using 23,422 individuals undergoing surgery for gallstones with 348,373 controls with no record of gallstones. 11 loci did not retain significance at the genome-wide threshold. Ten retained a suggestive association (*P* < 5 * 10^−5^; *ABO, ADAMTS20, FOXP1, GMDS-DT, GOT1*, chr18:rs542917327, *ITCH, NPAS2, SH3BP4, TBC1D32*) whilst one did not (*P* = 2.75 * 10^−4^; *LMNB1*). Direction and size of effect was very similar for all 55 loci suggesting detection bias had minimal influence on results. Full results at each locus are available in Supplementary Materials 4. A forest plot comparing effect sizes in shown in Supplementary Figure 6.

### Association of Lithogenic Loci with Phenotypic Traits

Assessment of serum lipids was undertaken for each of the 55 lithogenic variants from the UKB in all 377,000 individuals taken forward for GWAS. The lithogenic alleles were heterogeneous in their influence on serum lipids with some lithogenic alleles promoting hyperlipidaemia whilst others demonstrated a protective effect (see Figure 3). The lithogenic variants of *ABCG8* and *HNF4A* (rs11887534-C and rs1800961-T respectively) both resulted in reductions of cholesterol of 0.10mmol/L (95% CI 0.09-0.12). The lithogenic variants of *GCKR* and *ABO* demonstrated highly significant increases in serum cholesterol (rs1260326-C: 0.05mmol/L [95% CI 0.05-0.06]; and rs115478735-T: 0.07 [0.06-0.07]). Figures for the other serum biochemistry markers are available in Supplementary Figures 7-12. Numeric results are shown in Supplementary Materials 5.

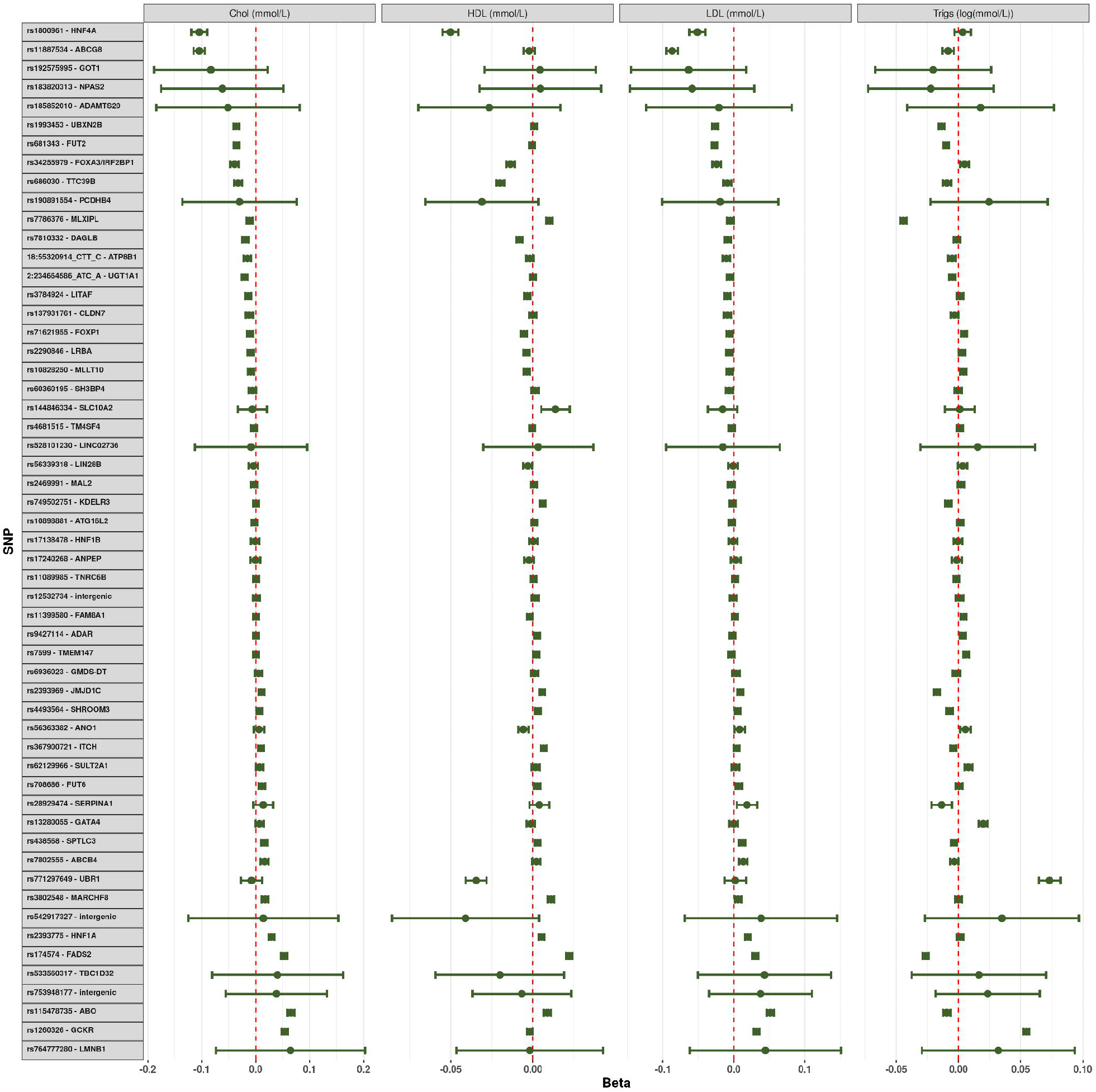

### Gene-Environment Interaction

Gene-environment interaction of SNPs with serum unconjugated bilirubin was undertaken at 54 of the lead SNPs from the main GWAS (excluding *UGT1A1*) in the 377,000 individuals. After Bonferroni correction for 54 analyses, a significance level of 0.00093 was determined. No variants reached statistical significance although nominal significance was seen in *LIN28B* (P=0.012), *ABCB4* (P=0.023), *JMJD1C*(P=0.008), *ANPEP*(P=0.041), *CLDN7*(P=0.011), *ATP8B1* (P=0.019) and *HNF4A* (P=0.002).

### Polygenic Risk Score

The PRS was significantly associated with gallstone disease in the validation cohort (maximum variance explained on the liability scale *R*^2^ = 2.5%, *P* = 2.62 * 10^−181^, when using a GWAS P value threshold of 0.0001). There was a 6-fold increase of risk of gallstones in those with the highest decile of genetic risk compared to those in the lowest with an approximately linear increase in risk with each increasing PRS decile (Figure 4). The PRS was strongly associated with increased BMI, waist circumference, hip circumference, liver enzymes (other than ALP), cystatin C and CRP (greater genetically-predicted risk of gallstones resulted in higher values for those traits). The PRS was associated with reductions in cholesterol, HDL, LDL, ApoB and ALP. Generally, for all traits with which the PRS was significantly associated there was an approximately linear increase (or decrease) with each increase in PRS decile and no apparent threshold effect. There were no obvious differences in levels of trigs, ApoA, LipoA, blood count markers, HbA1c or glucose (Supplementary Figures 13-15).

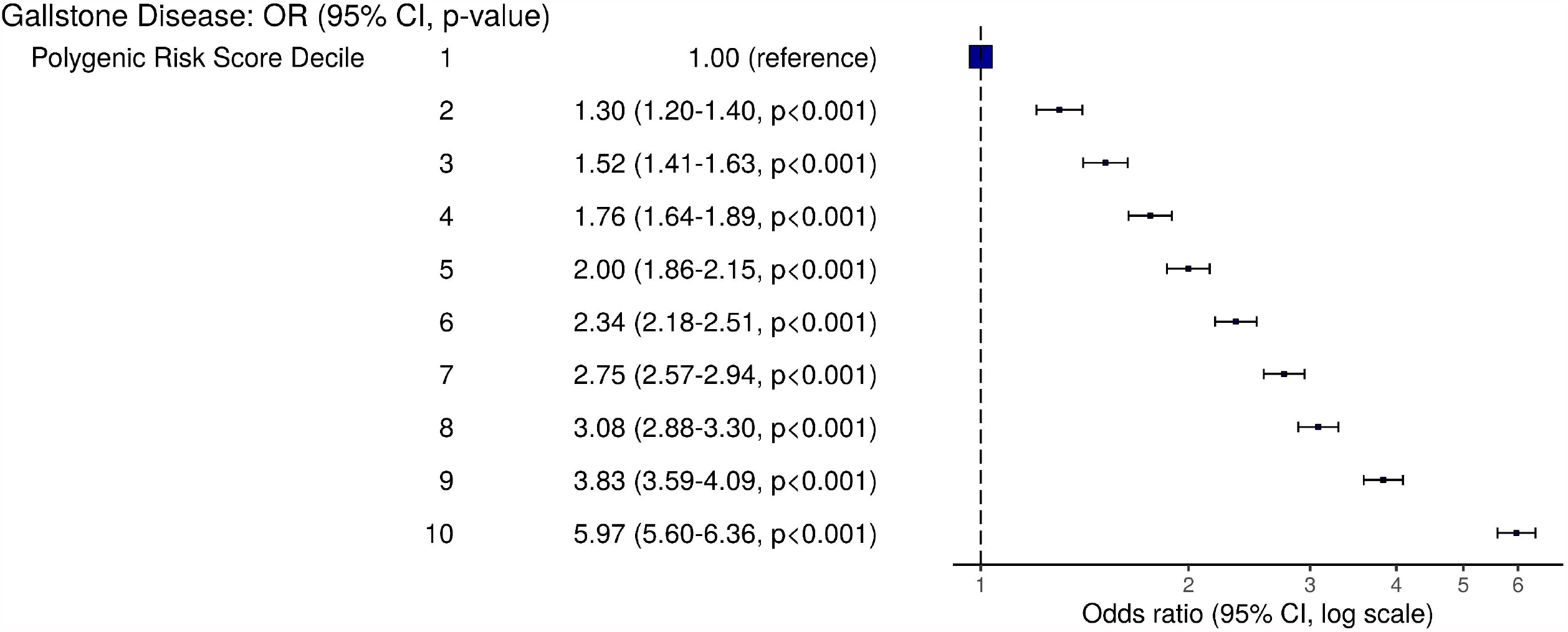

### Pathway Enrichment

172 genes showed significant association with gallstones (*P* < 2.59 * 10^−6^; Supplementary Materials 4) and were highly expressed in liver (*P* = 4.09 * 10^−6^; gallbladder expression was not available in MAGMA). 13 gene-sets were identified as significantly associated with gallstones (Table 3). Many of these gene-sets relate to glucoronidation, lipid metabolism and phospholipid transport. Additionally, Waldenström’s macroglobulinaemia and curated gene-sets of cancer drug resistance were identified which both contained *ABCB1* and *ABCB4*, both regulate biliary composition but are highly pleiotropic.

**Table 3:**
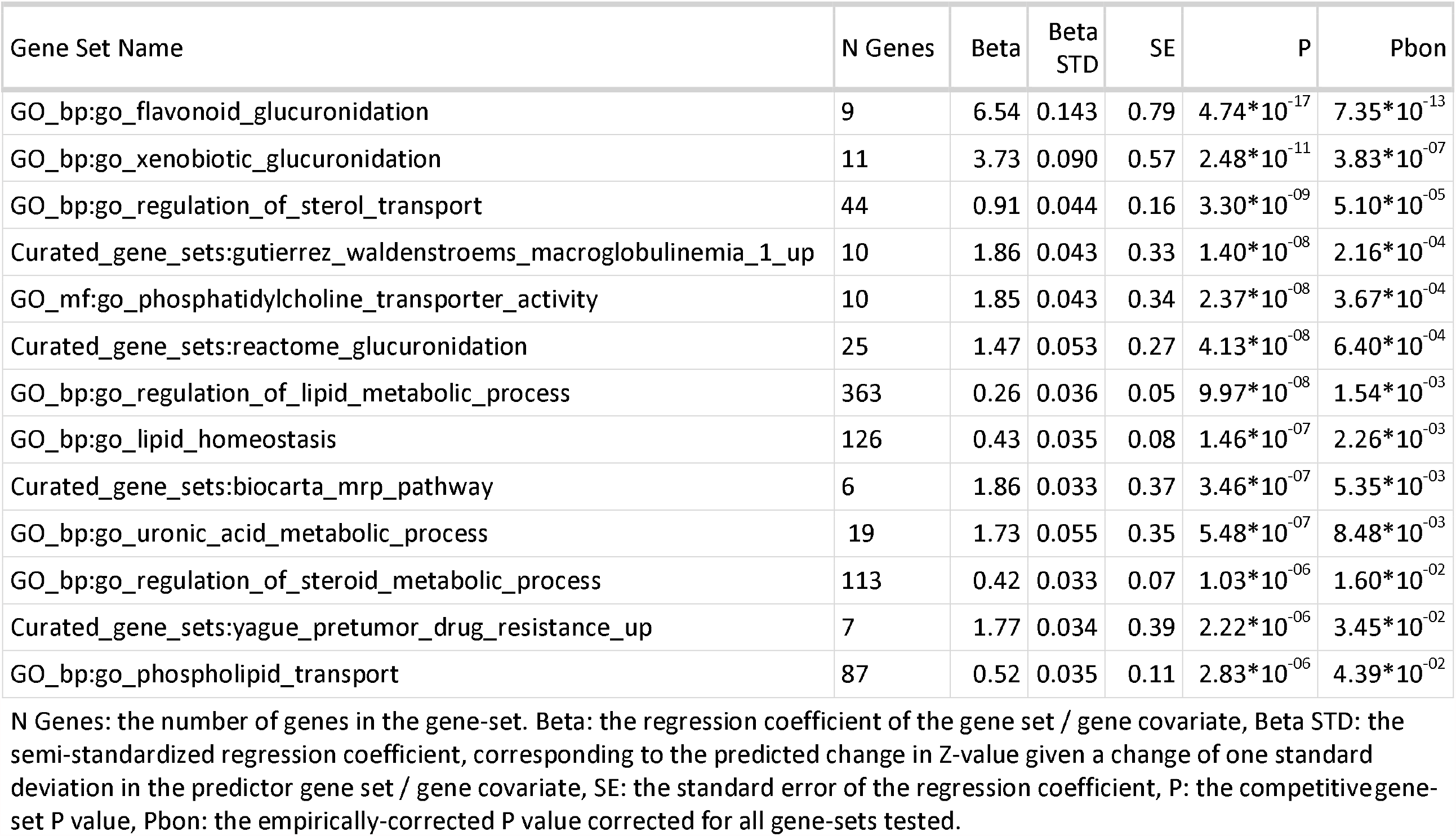
MAGMA Pathway Analysis - Significant Gene-Sets

Literature search identified two genes governing gastrointestinal motility (*ANO1* and *TMEM147*). Rs7599 lies within the 3’UTR region and the lithogenic allele (A) is associated with significantly reduced expression of *TMEM147* in the liver (*P* = 3.0 * 10^−47^). Data on effects of rs56363382 and all other significant SNPs in the *ANO1* loci were not available from GTEx. Both *ANO1* and *TMEM147* were found to be highly expressed in gallbladder mucosa in the Human Protein Atlas. 5 identified genes were involved in primary cilia function (*DYNC2LI1, TBC1D32, ADAMTS20, PKD2L1* and *POC1B*).

## Discussion

We report the largest and most detailed GWA of gallstones conducted worldwide with functional annotation of variants and gene-set analysis. We analysed 377,000 individuals (28,627 cases and 348,373 controls) from the UKB and replicated findings in a Scottish cohort (GS-SFHS) and a Finnish Cohort (FinnGen). We performed a genome-wide meta-analysis with UKB and FinnGen data. We identified 46 new lithogenic loci and replicated all 29 previously identified loci bringing the total number of identified gallstone-susceptibility loci to 75. Of the 75 loci, 12 were identified only in the UKB GWAS and not replicated or significant in the GWA meta-analysis. Through pathway enrichment we identified 13 gene-sets strongly associated with gallstones. Through FUMA annotation we identified two novel pathways that may influence gallstone development, gastrointestinal motility and ciliogenesis. Our results highlight important pathways that may be targeted by novel therapies and form the basis for further research into gallstones and other cholestatic disease.

Two newly identified loci are highly expressed in the gallbladder and govern gastrointestinal motility. *ANO1* (also known as *TMEM16A*) is a calcium-activated chloride channel widely expressed in the gastrointestinal tract with roles in automaticity and contraction of the smooth muscle cells via the interstitial cells of Cajal^33^. Administration of *ANO1* inhibitors (TMinh-23) in murine models impedes gastric emptying with improved oral but not parenteral glucose tolerance supporting the role of *ANO1* as a gastric motility promoter^33,34^. *ANO1* is also expressed in the biliary epithelium where it has another role in secretion of fluid from the apical cholangiocyte membrane in response to bile acids suggesting it may promote increased bile flow^35^. *ANO1* may influence gallstone formation via gallbladder contractility and biliary flow. Given the possible impact on gallbladder motility and biliary composition, we suggest that any trials of TMinh-23 assess for development of gallstones as an important safety consideration. *TMEM147* is known to inhibit transfer of M3 muscarinic receptors to the cell surface^36^. M3 receptors are highly expressed in smooth muscle cells of the gallbladder and may influence gallbladder contractility and biliary stasis^37^. The variants may also influence gallstone formation through increased colonic transit time which is also associated with gallstones and believed to alter enterohepatic bile acid circulation^14^.

Five loci relate to function of primary cilia. *TBC1D32* controls ciliogenesis within the neural tube and interacts with the sonic hedgehog protein which has wider roles in organogenesis and axonal guidance^38^. The metalloproteinase encoded by *ADAMTS20* also plays a pleiotropic role in ciliogenesis and hedgehog signaling with mutant variants causing ciliopathies^39^. *POC1B* is essential for formation of the centriole and ciliogenesis during cell division^40^. *PKD2L1* is involved in regulating calcium concentration within the primary cilia^41^ and disorders of calcium signaling of the primary cilia are thought to result in cholestasis^42^. Three independent signals within the *ABCG8* locus were intronic or 3’-UTR variants of *DYNC2LI1. DYNC2LI1* has a role in ciliogenesis with mutations causing severe ciliopathies^43^. Variants within the sonic hedgehog gene and *DYNC2LI1* have previously been associated with gallbladder cancer and gallstone formation^44^ whilst *TBC1D32* and *ADAMTS20* (novel loci, not replicated) and *PKD2L1* and *POC1B* (identified in the meta-analysis) are novel loci. We suggest that abnormalities in cholangiocyte ciliogenesis and ciliary function may contribute to biliary stasis with gallstone formation. Abnormal expression or function of cholangiocyte cilia causes other cholestatic disease and malignancies^45^. At present, therapies targeting primary cilia are limited. Ricolinostat, a selective inhibitor of histone deacetylase 6, is under investigation in several malignancies but its effect on gallstones remains unknown^45^.

MAGMA analysis identified thirteen gene-sets. Most were related to lipid homeostasis and glucoronidation. Gene-sets for Waldenström’s macroglobulinaemia and multidrug-resistance pathways were enriched with enrichment driven by *ABCB4* and *ABCB1. ABCB4* encodes a phosphatidylcholine flippase which effluxes of phosphatidylcholine into bile^46^. Phosphatidylcholine acts as a solvent for biliary cholesterol. *ABCB4* mutations are a cause of early-onset gallstones and low-phospholipid-associated cholelithiasis^47^. *ABCB1* is a neighbouring gene involved in phospholipid and drug excretion with broad specificity^46^. The pathway enrichments are most likely a feature of pleiotropy as these genes have several distinct functions. However, serum IgG and IgM extracted from patients with Waldenström’s macroglobulinaemia results in nucleation of biliary cholesterol, possibly representing an alternate mechanism directly linking gallstones with Waldenström’s macroglobulinaemia^48^.

Based on our results we suggest the following priorities for further research. 1. Direct genotyping at all 75 lead SNPs in individuals with corresponding cholecystectomy specimens for biochemical analysis of biliary content, gallbladder epithelial protein expression and cholangiocyte protein expression. 2. Assessment of gallbladder emptying with hepatobiliary scintigraphy in patients genotyped at *ANO1* and *TMEM147* to assess the influence of the lithogenic variants on gallbladder motility. 3. Assessment of gallstone risk in murine models with pharmacological modification of gallbladder motility (TMinh-23) or ciliotherapy (ricolinostat). 4. Validation of our findings in a large cohort in which radiological evaluation of gallstone disease has been undertaken.

The strengths of this study include a more complete definition of gallstones than in previous studies which reduces the rate of case misclassification. Notably, previous studies have identified up to 18,417 cases whilst we identify 28,627 from the UKB. We use three large cohorts with replication of the majority of novel findings. Many of the loci we identify have plausible relationships to gallstones which we explore in pathway analysis, tissue expression analyses and through assessing the impact of risk alleles on serum biomarkers. The limitations of this study include misclassification of individuals with asymptomatic gallstones that have not been detected clinically. Relying on clinical disease as the definition of exposure has been common to all earlier GWAS of gallstones although our selection of relevant codes has improved our sensitivity relative to other studies. The phenotypic characterisation of participants in the UKB is limited to less invasive procedures such as venepuncture and no data on biliary composition or pathological records of gallstone composition following cholecystectomy were available. This impedes the interpretation of cholesterol-influencing SNPs as SNPs may lead to reduced serum cholesterol through biliary excretion, increased serum cholesterol through intestinal absorption or a balanced combination of both with no net change in serum cholesterol. All three scenarios may lead to similar changes in biliary cholesterol yet demonstrate opposing effects on serum cholesterol. Finally, the definition of gallstones in the FinnGen cohort was not identical to the UKB as we used the published summary association statistics from the FinnGen GWAS although the majority of UKB cases would have met the definition used in FinnGen.

In summary, we have performed a GWAS of gallstone disease using UKB data, performed a meta-analysis with the FinnGen cohort and identified 46 novel variants associated with gallstone disease. We identify the first known gallstone-susceptibility variants that influence gastrointestinal motility, and we identify key priorities for research into gallstone formation and preventative treatments.

## Data Availability

Data from the individuals studies is available on request to verified authors.

## Acknowledgements

Generation Scotland received core support from the Chief Scientist Office of the Scottish Government Health Directorates [CZD/16/6] and the Scottish Funding Council [HR03006] and is currently supported by the Wellcome Trust [216767/Z/19/Z]. Genotyping of the GS:SFHS samples was carried out by the Genetics Core Laboratory at the Edinburgh Clinical Research Facility, University of Edinburgh, Scotland and was funded by the Medical Research Council UK and the Wellcome Trust (Wellcome Trust Strategic Award “STratifying Resilience and Depression Longitudinally” (STRADL) Reference 104036/Z/14/Z).

This work was funded by a Medical Research Council Clinical Research Training Fellowship awarded to CJF (MR/T008008/1). ADB acknowledges funding from the Wellcome PhD training fellowship for clinicians (204979/Z/16/Z), the Edinburgh Clinical Academic Track (ECAT) programme. CH and JFW acknowledges support from the MRC Human Genetics Unit programme grant, “Quantitative traits in health and disease” (U. MC_UU_00007/10). NCH is supported by a Wellcome Trust Senior Research Fellowship in Clinical Science (ref. 219542/Z/19/Z). NLM is supported by the British Heart Foundation through a Senior Clinical Research Fellowship (FS/16/14/32023) and a Research Excellence Award (RE/18/5/34216). RKS is supported by the Wellcome Trust (grant 210752/Z/18/Z). AS acknowledges support from the Academy of Medical Sciences/the Wellcome Trust/ the Government Department of Business, Energy and Industrial Strategy/the British Heart Foundation/Diabetes UK Springboard Award [SBF006\1109].

## Ethical Approval

The UKB received ethical approval (Research Ethics Committee reference number: 11/NW/0382). Ethical approval was granted for Generation Scotland: Scottish Family Health Study by NHS Tayside Research Ethics Committee (reference number: 05/S1401/89).

## Author Contributions

CJF conducted the genome-wide association study and drafted the manuscript. CJF, ADB, PKJ, DWC, PRHJT and JFW developed the code required for data analysis. CJF, TMD and JFW assisted with data access requests. RP, EMH, AI and JFW assisted with data storage and server maintenance. DJP, CH and AC contributed data from Generation Scotland. CJF, EMH, AS, SJW, CAS, TMD, RKS, NLM, CLMS and RP were involved in study design and conception. JAF, NCH, PR, NLM, RKS, SJW and EMH were involved in critical appraisal of the manuscript and placing the research in the biological context. ADB, AC, CH, AI, JFW and AS were involved in critical appraisal of the manuscript in relation to GWAS methodology. CJF, EMH, RKS, AS and AI were involved in pathway enrichment analyses.

