## Supplementary Materials 1 for "Genome-wide Analysis Identifies Novel Gallstone-susceptibility Loci Including Genes Regulating Gastrointestinal Motility"

Description of Supplementary Materials

### Supplementary Materials 1 – Supplementary Methods, Tables and Figures

- Methods
  - Establishment of Cohorts
  - Identification of gallstone disease
  - Genotyping and blood sampling
  - Analysis of P values below the limit of numerical precision
- Supplementary Tables
  - Supplementary Table 1: Diagnostic codes for gallstones
  - Supplementary Table 2: Diagnostic codes for surgical procedures for gallstones
  - Supplementary Table 3: Diagnostic codes for rare biliary pathology
  - Supplementary Table 4: Baseline Characteristics of Gallstone Cases and Controls in the UK Biobank
  - Supplementary Table 5: Replication of GWAS findings in the Generation Scotland: Scottish Family Health Study and FinnGen Cohort
- Supplementary Figures
  - Supplementary Figure 1: Rectangular format Manhattan plot for the GWA Meta-analysis
  - Supplementary Figure 2: Truncated rectangular format Manhattan plot for the GWA Meta-analysis
  - Supplementary Figure 3: Rectangular format Manhattan plot for the UKB GWAS
  - Supplementary Figure 4: Truncated rectangular format Manhattan plot for the UKB GWAS
  - Supplementary Figure 5: Quantile-quantile plot
  - Supplementary Figure 6: Comparison of effect sizes between main GWAS and the “Procedure-only” subgroup analysis (participants with gallstones who did not require a surgical procedure were excluded from this analysis)
  - Supplementary Figure 7: Association with Serum Lipid Particles
  - Supplementary Figure 8: Association with Serum Liver Enzymes
  - Supplementary Figure 9: Association with Serum Bilirubin Fractions
  - Supplementary Figure 10: Association with Serum Blood Count
  - Supplementary Figure 11: Association with Serum Glucose and HbA1c
  - Supplementary Figure 12: Association with Serum C-Reactive Protein and Cystatin C
  - Supplementary Figure 13: Polygenic Risk Score Association with Phenotypic Traits (Part 1)
  - Supplementary Figure 14: Polygenic Risk Score Association with Phenotypic Traits (Part 2)
  - Supplementary Figure 15: Polygenic Risk Score Association with Phenotypic Traits (Part 3)

### Supplementary Materials 2A - FUMA Parameters and Data Dictionary

- Data dictionary for each file in the main FUMA results
- List of parameters passed to the Functional Mapping and Annotation of GWAS results

### Supplementary Materials 2B - FUMA Results

- FUMA results in xlsx file (each sheet of the file is a specific FUMA output file as described in 2A)

### Supplementary Materials 3 - STREGA Checklist

- STREGA checklist with page references to submitted version

### Supplementary Materials 4 - Significant GWAS Results

- xlsx file with main significant SNPs for main GWAS and subgroup analyses, sheets of xlsx file as follows:
  - “Gallstone Disease versus Controls”: SNPtest output for all SNPs reaching genome-wide significance ($P<5*{10}^{-8}$) in main GWAS
  - “Conditional Analyses Results”: Results for each of the conditional analyses (adjusting for each new lead SNP until no significant results remain)
  - “Lead 55 SNPs Main GWAS”: Snptest output for the 55 SNPs identified as independent lead SNPs in the main GWAS
  - “Procedure-Only Sens. Analysis”: SNPtest output for each of the 55 SNPs identified in the main GWAS after exclusion of individuals with a history of gallstones but not cholecystectomy or other gallstone extraction procedure
  - “GWA Meta-Analysis”: METAL output for all SNPs reaching genome-wide significance ($P<5*{10}^{-8}$) in GWA meta-analysis
  - “MAGMA Genes”: MAGMA output for all genes reaching gene-based significant in gene-based analysis

### Supplementary Materials 5 - Biomarker Regression Results

- Numeric results from each of the biomarker lattice plots covering linear regression coefficients - xlsx file which contains one sheet per biomarker (total cholesterol, low-density lipoprotein, triglycerides, high-density lipoprotein, apolipoprotein A, apolipoprotein B, lipoprotein A, alkaline phosphatase, alanine aminotransferase, aspartate aminotransferase, gamma glutamyltransferase, unconjugated bilirubin, conjugated bilirubin, total bilirubin, haemoglobin, red cell count, reticulocyte percentage, glucose, glycated haemoglobin, C-reactive protein, cystatin C)
