## Supplementary material for "Genome-wide Analysis Identifies Novel Gallstone-susceptibility Loci Including Genes Regulating Gastrointestinal Motility": Figure

Figure 1: Flowchart describing participant recruitment in UK Biobank, Generation Scotland: Scottish Family Health Study and FinnGen

Figure 2: Circular Manhattan plot for the association of genetic variants with gallstone disease (43,639 cases and 506,798 controls). Each variant is plotted based on chromosome and position on the X-axis and -log10 P values on the Y-axis. The dashed red line represents genome-wide significance ($P=5*{10}^{-8}$). Variants highlighted in bold were significant on GWA meta-analysis. Other variants were only significant in the UK Biobank cohort. *P value at ABCG8 locus has been truncated and is approximately $2.21*{10}^{-715}$.

Figure 3: Impact of each lithogenic allele on the measured serum cholesterol fractions. Each point represents the beta-coefficient from an age- and sex-adjusted linear regression and the error bar represents the 95% confidence interval. Triglycerides were log-transformed prior to analysis. Chol - Total cholesterol; HDL - high-density lipoprotein; LDL - low-density lipoprotein; trigs - triglycerides.

Figure 4: Odds ratio plot demonstrating odds of gallstones by decile of the polygenic risk score. Each decile of the risk score is compared to the group with the lowest risk of gallstones (Decile 1).
