## Supplementary Materials 1 for "Genome-wide Analysis Identifies Novel Gallstone-susceptibility Loci Including Genes Regulating Gastrointestinal Motility"

### Supplementary Materials 1: Methods, Tables and Figures

### Establishment of Cohorts

#### UK Biobank

The UKB is a prospective cohort study with 502,616 participants recruited between 2006 and 2010. Participants were men and women aged between 37 and 73 at time of recruitment. Initial assessment involved self-completed touch-screen questionnaire, computer-assisted interview, physical and functional measures and collection of blood samples. Additional data has been generated from health record linkage to national registries and hospital discharges and genotyping of participants. Over 2.5 million hospital admissions were available for analysis with data available on diagnosis, operative procedures and admission dates. Additional primary care records were made available for 270,000 participants.

The recruitment process, consent process and data collection has been described extensively elsewhere^1–3^. Data is available to researchers after a two-stage online application process. The UKB received ethical approval (research ethical committee reference 11/NW/0382). UKB data access was approved under projects 30439 (phenotype data) and 19655 (genotype data).

#### Generation Scotland: Scottish Family Health Study

Generation Scotland: Scottish Family Health Study (GS-SFHS) is a family-based genetic epidemiology study with DNA and socio-demographic and clinical data covering 24,096 volunteers across Scotland aged 18-98 years, from 2006 to 2011. Participants were identified within general practices in Scotland and were aged between 35 and 55 years with at least one first degree relative aged 18 years or over and at least one full sibling group. Participants underwent baseline interview with storage of blood samples for genotyping. Records for individuals were also linked to hospital- and community-based records using their community hospital index number provided to all citizens in Scotland registered with a general practice. A full description of the GS-SFHS protocol has been published previously^4,5^. Ethical approval was granted by NHS Tayside Research Ethics Committee (REC reference number 05/S1401/89).

#### FinnGen

FinnGen combines several Finnish Biobanks with genotyping data linked to digitised national health registries. The researchers have conducted GWAS for ICD10 categories and summary association statistics from Data Release 4, including 176,899 participants, are now publicly available^6^. The FinnGen study protocol was approved by the Ethical Review Board of the Hospital District of Helsinki and Uusimaa (Nr HUS/990/2017).

### Identification of Gallstone Disease

#### UK Biobank

Gallstone disease was defined as any hospital admission with an ICD-9 or ICD-10 code relating to gallstones, any primary care encounter with a Read code relating to gallstones, any self-reported history of gallstones at study enrolment and any procedure specifically due to or highly likely to be due to gallstones. Cholecystectomy performed in a patient without a specific code relating to gallstones was considered as gallstone disease except when a reasonable alternative indication (gallbladder polyps, choledochal cyst, trauma, benign or malignant hepatobiliary pathology etc.) was present. Gallstone disease is the largest indication for cholecystectomy with under 2% performed for alternative indications^7^. After exclusions for alternative pathology, the risk of falsely classifying a patient as having gallstones after undergoing cholecystectomy is extremely small. This risk is also vastly outweighed by the risk of misclassifying participants as controls after undergoing cholecystectomy as most of these cases will be due to gallstones. Procedures which specifically mention gallstones were considered as cases of gallstones irrespective of other diagnoses. A full list of the relevant ICD, OPCS, Read and UK-Biobank-specific codes are available in Supplementary Tables 1-3.

#### Generation Scotland: Scottish Family Health Study

Gallstone disease was defined as any hospital admission or primary care consultation resulting in generation of an ICD, OPCS or Read code relating to gallstones or treatment of gallstones. The same exclusions were applied for treatment such as cholecystectomy where a relevant alternative indication was also known. Participants in GS-SFHS were not asked to report a history of gallstone disease at study onset.

#### FinnGen

Participants in FinnGen were linked to several national registries including the Care Register for Healthcare and the Register of Primary Healthcare Visits. These registries spanning decades were linked to genotype data using unique national personal identification numbers assigned to all Finnish citizens and residents. The most similar overall definition to that used in the UK Biobank was ICD10 “K80” and ICD9 “574” (cholelithiasis). A closer definition of “K80” and/or “K81” (cholelithiasis and cholecystitis) was not available with the public data download. A much broader definition included cholangitis, pancreatitis (acute and chronic), other biliary disease and other pancreatic disease but this was thought to be too broad a definition for inclusion in meta-analysis.

### Genotyping and Blood Sampling

#### UK Biobank

For the UKB, collection of blood for genotyping was performed at study enrolment for approximately 470,000 individuals, the remainder were excluded due to insufficient sample volume. The cardiorespiratory-focused Affymetrix UK BiLEVE Axiom array was used in 50,000 participants and the Affymetrix UKB Axiom array was used for the remaining participants, the arrays were over 95% similar. Genotyping was performed in 106 batches (4000-5000 individuals per batch). Approximately 900,000 SNPs were directly called with imputation resulting in 93 million SNPs for assessment. The full Affymetrix protocol and description of quality control performed prior to release of results to researchers have been published previously^1,8,9^.

#### Generation Scotland: Scottish Family Health Study

For GS-SFHS, collection of blood samples and subsequent genotyping was performed at study enrolment with 20,032 having DNA extracted via either blood or saliva. Genotyping was performed on an Illumina HiScan platform and genotypes were called automatically using GenomeStudio Analysis software v2011.1. Genotypes were imputed using the Haplotype Reference Consortium reference panel (HRC.r1-1) via the Sanger Imputation Server pipeline (<https://imputation.sanger.ac.uk>). A total of 24,161,581 SNPs were available for analysis. Genotype data covering each of the identified loci in the UK Biobank GWAS were extracted from GS-SFHS.

#### FinnGen

For FinnGen, individuals were genotyped with Illumina and Affymetrix arrays. Genotype data were imputed using the population-specific SISu v3 imputation reference panel of 3,775 whole genomes. After imputation 16,962,023 variants for 176,899 participants were available after imputation. A GWAS was performed using SAIGE^10^ with the following covariates: sex, age, 10 genetic principal components and genotyping batch.

### Analysis of P Values Below the Limit of Numerical Precision

In the analysis of chromosome 2, a region associated strongly with gallstone disease generated a total of 24 P values with a value recorded as 0. The variants had almost identical $\beta$-coefficients with the same direction of effect. The rs11887534 ABCG8 D19H variant is a known protein-altering variant and previously identified as the lead SNP by other GWAS. This variant was considered to be the lead SNP for this locus.

As P values cannot be exactly 0, a decision was made to set this value to $4.94*{10}^{-324}$ which is the “Unit in the last place” on a Linux 64-bit operating system). In reality the P value may be smaller than this but smaller values cannot typically be stored accurately by a 64-bit operating system.

The following evidence was considered in support of this approach:

- All 0-value P-values occurred within a small region of chromosome 2 (< 36KB range)
- All confidence intervals were relatively narrow and of similar margins
  - Lower 95% CI ranged from 1.79-1.96
  - Upper 95% CI ranged from 1.92-2.07
  - All 95% CI ranges were 0.11-0.13
- The nearest non-0 P-value in terms of magnitude lay within the same region (within the 36KB range) and was very small ($2.1*{10}^{-}262$)
- There are several very small P-values ($P<5*{10}^{-}100$) in the same region
- The smallest 203 SNPs (across all chromosomes) all lie on chromosome 2 within an 810KB region (relative to a 243,178KB range for the whole chromosome)
- Several studies already published have revealed very small P values for the suspected lead SNP (rs11887534) in this region
  - A meta-analysis of GWA and candidate gene studies demonstrated a P-value of $2.3*{10}^{-}187$ for the same variant^11^
  - A GWAS published using Global Biobank Engine (GBE) data (based on 3-level ICD10 codes using UK Biobank data) found a P-value of $2.93*{10}^{-}298$ for the same SNP^12^
  - The GBE papers (as discussed in the main manuscript) relied on less sensitive phenotypic characterisation of gallstone disease and as such will have considered some true gallstone cases as controls thus reducing the apparent effect at true causative loci
  - Both the meta-analysis and GBE data reveal similar odds ratios (OR 2.42 95% CI[2.29-2.57] and OR 2.01 95% CI[1.94-2.08] respectively) to this analysis (OR 2.01 95% CI[1.96-2.07])
- All of the 0-value P values in this region are 0 (stored as 0.000000e+00) and are not missing or NULL, this is a known behaviour of SNPtest that extreme effect sizes will generate a 0-value P value and not a missing value
- Each of the 24 0-value SNPs were marked in the SNPtest output “comment” variable as “igamc_underflow_error”, this is a known flag generated when attempting to generate an extremely small P value from a chi-squared test and can represent either extreme effect sizes or an error with the data (<https://mathgen.stats.ox.ac.uk/genetics_software/snptest/old/snptest_v2.3.0.html>)
  - In the context of the other evidence discussed and given there were no other errors with the data and these SNPs do not generate errors when run for other phenotypes, it was thought that this represents a genuine P value less than the Unit in the Last place (ULP) rather than an error with the data
- The Unit in the Last Place (ULP; <https://en.wikipedia.org/wiki/Unit_in_the_last_place>)^13^ is the smallest possible value that can be stored by a computer system and is close to the expected values for the data
  - For 64-bit Linux systems the smallest finite value (other than 0 itself) that can be stored is thought to be in the region of $1*{10}^{-}308$ <https://www.sciencedirect.com/science/article/pii/S0010448597000869> (it is not possible to store a value smaller than the ULP other than 0)
  - Experimenting within two distinct 64-bit Linux systems (one of which was used for the GWAS), the smallest number possible in both cases was $4.940656e-324$ (identical in both systems)
  - Attempting to divide or subtract from this number results in either the same number or a 0 value
  - Attempting to enter a value smaller than this number returns either $4.940656e-324$ or 0 and never a value in between
  - Entering $3e-324$ gives $4.940656e-324$ whilst $2e-324$ gives 0 suggesting any value between 0 and $4.940656e-324$ is not possible
  - Attempting to multiply the number by a small quantity (e.g. 1.1) does not change the value whilst multiplying by a 1.5 causes the value to double in-keeping with this number being the smallest division possible within the operating system
- Finally in support of this approach and also of interest is another related GWAS of serum bilirubin which may have suffered from a similar problem^14^
  - The paper states: “There were four most significant UGT1A1 SNPs in the proximal promoter region and intron 1, physically close to the UGT1A1 TATA box polymorphism, all of which showed similar significant signals with nearly identical b coefficients (rs6742078, rs887829, rs4148324, rs4148325).”
  - In the combined meta-analysis component of the paper the P value for these SNPs is listed as: $P<5*{10}^{-324}$

As plotting the P value on a standard Manhattan plot is not possible when it is 0 (this would be at infinity on the scale) each of the 0-value P values were changed to the ULP for the purposes of plotting. Given the computational complexity of moving beyond a 64-bit system, no 128-bit system is routinely available for conducting this analysis. It is therefore not possible to confirm the hypothesis by seeking greater precision in this way.

### Supplementary Tables

1. Supplementary Table 1: Disease Codes: Gallstone Pathology
2. Supplementary Table 2: Disease Codes: Rare Biliary Pathology
3. Supplementary Table 3: Operation Codes: Gallstone-Related Procedures
4. Supplementary Table 4: Baseline Characteristics of Gallstone Cases and Controls in the UK Biobank
5. Supplementary Table 5: Replication of GWAS Findings in the Generation Scotland: Scottish Family Health Study and FinnGen Cohort

| Supplementary Table 1: Gallstone-Related Pathology: Disease Codes | | | | | |
| --- | --- | --- | --- | --- | --- |
| Diagnosis | ICD10 | ICD9 | UKB Code | READ V2 Code | Additional READ V3 Codes*^1^* |
| Cholelithiasis | K80 | 574 | 1162 | J642., J6420-J6422, J642z, J64z., J64z0, J64z1, J64zz, J6705, Jyu80 | 0.1965, 1965, X308A, X308I, X75rk, XaDc4, XE0dL |
| Cholecystitis | K81 | 575.0, 575.1 | 1163 | J640., J6400-J6401, J640z, J641., J6410-J6411, J641z, J650., J6500-J6504, J650z, J651., J6510, J651y, J651z, J653., Jyu81 | X3089, X308B, XaDc4, XaFr2, XE0bF |
| Bile Duct Stone | K80.4 - K80.7 | 574.3 - 574.9 | 1160 | J643., J6430-J6431, J643z, J644., J6440-J6441, J644z, J645., J6450-J6452, J645z, J646., J67y1 | X308J, Xa6bl, XaAzY, XaE6v, XE0bD, XE0bE |
| Biliary Acute Pancreatitis | K85.1 | -*^2^* | - | J6707, J6711 | X308i, X308u, X309K, XaYaK |
| Post-cholecystectomy Syndrome | K91.5 | 576.0 | - | J660. | - |
| Retained Cholelithiasis following Cholecystectomy | K91.86 | 997.41 | - | - | X308K |
| Gallstone Ileus | K56.3 | 560.31 | - | J5030 | - |
| Other Gallstone code | - | - | 1161 | 4G2Z. | 0.4775, 4775, X79cS, X7ABd, XaaBJ |

| Supplementary Table 2: Gallstone-Related Procedures: Surgical Codes | | | | | |
| --- | --- | --- | --- | --- | --- |
| Procedure | OPCS4 | OPCS3*^1^* | UKB Code*^1^* | READ V2 Code | Additional READ V3 Codes*^2^* |
| **Cholecystectomy:** |  |  |  |  |  |
| Excision of gallbladder*^3^* | J18 | 522 | 145.5 | .79O1, X20bU, X20bV, X20bW, Xa9Zu, XaBlj, XacY3, XaQkQ, XE0E6-XE0E7 | 14ND., 78100-78106, 7810y, 7810z |
| **Other Gallbladder Procedures:** | | | |  |  |
| Removal of calculus from gallbladder | J21.1 | 521 | - | X20bc, X20bd, XE0EA, XM1IM | 78130 |
| Percutaneous fragmentation of calculus in gallbladder | J24.2 | - | - | X20be | 78151 |
| Percutaneous dissolution therapy of calculus in gallbladder | J24.3 | - | - | XM0Bz | 78152 |
| Extracorporeal fragmentation of calculus in gallbladder | J26.1 | - | - | X20cq | 78170 |
| **Biliary Tree Procedures:** |  |  |  |  |  |
| Open removal of calculus from bile duct and drainage of bile duct | J33.1 | 511 | - | - | 78260 |
| Open removal of calculus from bile duct NEC | J33.2 | 511 | - | X20c1-X20c4, X70aa, XE0EJ | 78261 |
| Endoscopic retrograde extraction of calculus from bile duct | J38.1, J41.1 | - | - | XE0EL | 782B0 |
| Endoscopic retrograde lithotripsy of calculus of bile duct | J41.3 | - | - | XaLeI | - |
| Removal of calculus from bile duct along T tube track | J49.1 | - | - | - | 782N0, 782N1 |
| Percutaneous removal of calculus from bile duct along T tube track | J49.2 | - | - | - | - |
| Extracorporeal lithotripsy of calculus in bile duct | J52.1 | - | - | - | 782Q0 |
| Removal of calculus from pancreatic duct | J42.3, J60.2 | 530.2 | - | X20aq, Xa3uL | 782F2, 78361 |
| Removal of calculus from hepatic duct | - | - | - | Xa7ew | 78041, 78060 |
| Extracorporeal shockwave lithotripsy of calculus of pancreas | J68.1 | - | - | XaMlf | 783F0 |
| Percutaneous transhepatic removal of calculus from bile duct | J76.1 | - | - | XaMr7 | 78091, 782T0 |
| **Miscellaneous:** |  |  |  |  |  |
| Gallstones removed (other) | All codes combined | All codes combined | 152.8 | .4G9A, XaB9c, XaEWK, XaFB5, XM16o | 76487 |

| Supplementary Table 3: Rare Biliary Pathology: Disease Codes | | | | | |
| --- | --- | --- | --- | --- | --- |
| Diagnosis | ICD10 | ICD9 | UKB Code | READ V2 Code | Additional READ V3 Codes*^1^* |
| **Malignant Diseases of:** | | | | | |
| Gallbladder | C23 | 156 | 1025 | .B163, X78Fb, X78gY, X78P7, XaFrF, XaFrr, XE1vY, XE1xt | B160., B16y., B16z. |
| Liver and Intra-hepatic Bile Duct | C22 | 155 | 1024 | .B161, X77nk, X78Oz, X78P0, Xa97q-Xa97r, XE1xn, XE1xp, XE2xA, XM1FE | B150., B1500-B1503, B150z, B151., B1511-B1514, B151z, B152., B15z., BB5D8, BB5Dz |
| Duodenum | C17.0 | 152.0 | - | X78NL | B120. |
| Unspecified Biliary Tract | C24 | 156 | - | X78Fc-X78Fd, X78gb, X78PC, X78PX, XaDbr, XaFr0, XE1xv | B161., B1610-B1613, B161z, B162., B163., BB5D1, BB5D7 |
| Pancreas | C25 | 157 | 1026 | .B17., .B17Z, X309B, X309D, X78gd, X78Pg-X78Ph, XaFrH, XaFrp, XE1y5 | B170.-B175., B17y., B17yz, B17z. |
| **Carcinoma in Situ (or Neoplasm of uncertain behaviour) of:** | | | | | |
| Liver, Gallbladder,  Bile Duct or Duodenum | D01.5 | 230.8 | - | X3080, X308Y, X78mC, X78PA, X78PD, X78PF | B808., B8080-B8087, B808z, B903., B9030-B9037, B903z |
| Duodenum | - | - | - | - | B8070, B9021 |
| Pancreas | - | - | - | - | B80z0, B9051 |
| **Benign Neoplasm of:** |  |  |  |  |  |
| Gallbladder | - | - | - | - | B1752-B1753 |
| Duodenum | D13.2 | 211.2 | - | XE1vv | B7120 |
| Liver | D13.4 | 211.5 | - | Xa0lo | B1750, B1751, B1754, B7158 |
| Biliary Tract | D13.5 | 211.5 | - | X78oB, X78oD, X78oG, X78PE, Xa0EM | B175., B1755-B1757, B175z |
| Pancreas | D13.6 | 211.6 | - | X309A, X309E, X78oE, X78Pi-X78Pj, Xa0EL, Xa98M-Xa98N, XaJgM, XE1vz, XM0B4, XM1G8 | B716., B7160-B7163, B716z, B717., B7170, B717z |
| Endocrine Pancreas | D13.7 | 211.7 | - | X50GW, X78cy, X78cz, X78d2-X78d3 | - |
| **Injury of:** |  |  |  |  |  |
| Liver, Gallbladder or Bile Duct | S36.1 | 864, 868.02, 868.12 | - | X307D, XA045, XA07B, XA07F, XA07H-XA07O, XA07Q-XA07Z, Xa25g-Xa25i, Xa25m, Xa25o, XE1mD, XE1mI-XE1mJ | S740., S7400-S7404, S740y, S741., S7410-S7141, S741y, S741z, S74z., S7802, S7812, SB222 |
| **Other:** |  |  |  |  |  |
| Gallbladder polyp or cholesterolosis | K82.4 | 575.6 | - | XaIIr | J656., J66y2, PB6y0 |
| Choledochal Cyst | K83.5 | -*^2^* | - | X20bh | - |

| Supplementary Table 4: Baseline Characteristics of the UK Biobank Gallstones GWAS cohort | | | | |
| --- | --- | --- | --- | --- |
|  |  | Gallstones | Controls | p |
| Sex | Female | 20670 (70.9) | 181607 (52.2) | <0.001 |
|  | Male | 8473 (29.1) | 166250 (47.8) |  |
| Age at Recruitment | Mean (SD) | 59.2 (7.3) | 56.8 (8.0) | <0.001 |
| Weight (kg) | Mean (SD) | 81.6 (16.8) | 78.1 (15.8) | <0.001 |
| BMI (kg/m2) | Mean (SD) | 29.6 (5.5) | 27.2 (4.6) | <0.001 |
| Systolic Blood Pressure (mmHg) | Mean (SD) | 139.2 (18.4) | 138.3 (18.6) | <0.001 |
| Diastolic Blood Pressure (mmHg) | Mean (SD) | 82.5 (9.9) | 82.3 (10.1) | 0.043 |
| Alanine Aminotransferase (U/L) | Mean (SD) | 25.1 (17.0) | 23.4 (13.8) | <0.001 |
| Aspartate Aminotransferase (U/L) | Mean (SD) | 26.8 (13.0) | 26.2 (10.3) | <0.001 |
| Alkaline Phosphatase (U/L) | Mean (SD) | 91.5 (36.2) | 83.1 (25.5) | <0.001 |
| Gamma Glutamyltransferase (U/L) | Mean (SD) | 44.8 (57.7) | 36.9 (40.4) | <0.001 |
| Bilirubin (Direct; μmol/L) | Mean (SD) | 1.8 (1.0) | 1.8 (0.8) | 0.098 |
| Bilirubin (Total; μmol/L) | Mean (SD) | 8.9 (4.8) | 9.2 (4.4) | <0.001 |
| Cholesterol (mmol/L) | Mean (SD) | 5.6 (1.2) | 5.7 (1.1) | <0.001 |
| Triglycerides (mmol/L) | Mean (SD) | 1.9 (1.0) | 1.7 (1.0) | <0.001 |
| Low-Density Lipoprotein (mmol/L) | Mean (SD) | 3.5 (0.9) | 3.6 (0.9) | <0.001 |
| High-Density Lipoprotein (mmol/L) | Mean (SD) | 1.4 (0.4) | 1.5 (0.4) | <0.001 |
| Apolipoprotein A (g/L) | Mean (SD) | 1.5 (0.3) | 1.5 (0.3) | <0.001 |
| Apolipoprotein B (g/L) | Mean (SD) | 1.0 (0.2) | 1.0 (0.2) | 0.006 |
| Lipoprotein A (nmol/l) | Mean (SD) | 43.8 (48.9) | 44.1 (49.5) | 0.293 |
| C-Reactive Protein (mg/L) | Mean (SD) | 3.7 (5.3) | 2.5 (4.3) | <0.001 |
| Glucose (mmol/L) | Mean (SD) | 5.3 (1.5) | 5.1 (1.2) | <0.001 |
| Glycated Haemoglobin (HbA1c; mmol/mol) | Mean (SD) | 37.3 (8.4) | 35.9 (6.3) | <0.001 |
| All Diabetes*^1^* | Yes | 4233 (14.5) | 26007 (7.5) | <0.001 |
|  | No | 24910 (85.5) | 321850 (92.5) |  |
| Type 2 Diabetes | Yes | 3898 (13.4) | 22264 (6.4) | <0.001 |
|  | No | 25245 (86.6) | 325593 (93.6) |  |
| *^1^*'All Diabetes' includes Type 1, Type 2, other types and unspecified diabetes mellitus. | | | | |
| Categorical data tested with Chi-squared test. Continuous data tested with Welch two-sample T-test. | | | | |

| Supplementary Table 5: Replication of GWAS Findings in the Generation Scotland: Scottish Family Health Study and FinnGen Cohort | | | | | | | | | | | | | | |
| --- | --- | --- | --- | --- | --- | --- | --- | --- | --- | --- | --- | --- | --- | --- |
| UKB | | | | | GS-SFHS | | | | | Finngen | | | | |
| RSID | Reference/ Effect Allele | OR (95% CI) | P | EAF | RSID | Reference/ Effect Allele | OR (95% CI) | P | r^2^ | RSID | Reference/ Effect Allele | OR (95% CI) | P | r^2^ |
| rs7802555 | A/C | 0.85 (0.83-0.88) | 1.37*10^-34^ | 0.139 | rs7802555 | A/C | 0.83 (0.73-0.95) | 4.77*10^-03^ | - | rs7802555 | A/C | 0.83 (0.81-0.84) | 4.03*10^-22^ | - |
| rs11887534 | G/C | 2.01 (1.96-2.07) | 4.94*10^-324^ | 0.066 | rs11887534 | G/C | 1.93 (1.67-2.24) | 3.98*10^-19^ | - | rs11887534 | G/C | 2.29 (2.23-2.34) | 2.49*10^-236^ | - |
| rs115478735 | A/T | 1.05 (1.03-1.08) | 2.00*10^-08^ | 0.186 | rs2519093 | C/T | 1.01 (0.89-1.15) | 3.49*10^-01^ | 0.98 | rs115478735 | A/T | 1.10 (1.08-1.11) | 5.05*10^-08^ | - |
| rs185852010 | C/T | 1.24 (0.99-1.55) | 2.77*10^-08^ | 0.001 | rs185852010 | C/T | 1.11 (0.32-3.89) | 9.77*10^-01^ | - | - | - | - | - | - |
| rs9427114 | T/C | 1.05 (1.04-1.07) | 1.47*10^-10^ | 0.490 | rs9427114 | T/C | 0.92 (0.84-1.01) | 5.72*10^-02^ | - | rs9427114 | T/C | 1.05 (1.03-1.06) | 9.08*10^-04^ | - |
| rs56363382 | C/T | 1.13 (1.09-1.16) | 6.51*10^-15^ | 0.080 | rs55953866 | C/T | 1.02 (0.86-1.21) | 9.19*10^-01^ | 0.88 | rs56363382 | C/T | 1.09 (1.06-1.12) | 5.77*10^-04^ | - |
| rs17240268 | G/A | 0.89 (0.86-0.92) | 3.03*10^-13^ | 0.090 | rs17240268 | G/A | 0.93 (0.79-1.09) | 3.63*10^-01^ | - | rs17240268 | G/A | 0.90 (0.88-0.93) | 1.25*10^-04^ | - |
| rs10898881 | A/G | 0.95 (0.94-0.97) | 1.88*10^-08^ | 0.453 | rs10898881 | A/G | 0.94 (0.86-1.03) | 1.53*10^-01^ | - | rs10898881 | A/G | 0.99 (0.97-1.00) | 3.02*10^-01^ | - |
| 18:55320914_CTT_C | CTT/C | 0.94 (0.92-0.96) | 4.63*10^-08^ | 0.181 | - | - | - | - | - | rs7226404 | A/C | 0.94 (0.92-0.96) | 3.80*10^-04^ | 0.66 |
| rs137931761 | C/CT | 0.94 (0.91-0.96) | 1.01*10^-08^ | 0.174 | rs17710 | A/T | 0.98 (0.86-1.13) | 7.57*10^-01^ | 0.73 | rs137931761 | C/CT | 1.00 (0.98-1.01) | 8.99*10^-01^ | - |
| rs1993453 | A/G | 0.91 (0.89-0.92) | 1.62*10^-29^ | 0.337 | rs1993453 | A/G | 0.84 (0.77-0.93) | 5.71*10^-04^ | - | rs1993453 | A/G | 0.88 (0.87-0.89) | 1.53*10^-20^ | - |
| rs7810332 | G/C | 0.94 (0.92-0.96) | 2.20*10^-09^ | 0.262 | rs7810332 | G/C | 0.94 (0.85-1.04) | 4.01*10^-01^ | - | rs7810332 | G/C | 0.95 (0.93-0.96) | 2.24*10^-04^ | - |
| rs174574 | A/C | 0.94 (0.93-0.96) | 3.04*10^-10^ | 0.353 | rs174574 | A/C | 0.96 (0.88-1.06) | 5.55*10^-01^ | - | rs174574 | A/C | 0.93 (0.92-0.95) | 3.39*10^-07^ | - |
| rs11399580 | G/GT | 0.95 (0.93-0.96) | 1.95*10^-10^ | 0.453 | rs11753865 | A/G | 0.99 (0.91-1.09) | 5.99*10^-01^ | 0.81 | rs9396784 | G/A | 0.94 (0.93-0.95) | 1.22*10^-05^ | 0.85 |
| rs71621955 | A/AT | 0.95 (0.94-0.97) | 1.08*10^-08^ | 0.433 | rs704251 | A/T | 0.97 (0.88-1.06) | 6.03*10^-01^ | 0.74 | - | - | - | - | - |
| rs681343 | C/T | 1.12 (1.10-1.14) | 2.76*10^-32^ | 0.492 | rs681343 | C/T | 1.08 (0.98-1.18) | 2.00*10^-01^ | - | rs681343 | C/T | 1.04 (1.03-1.05) | 3.97*10^-03^ | - |
| rs708686 | C/T | 1.06 (1.04-1.08) | 4.18*10^-12^ | 0.267 | rs708686 | C/T | 0.98 (0.88-1.09) | 7.90*10^-01^ | - | rs708686 | C/T | 1.10 (1.08-1.11) | 3.00*10^-11^ | - |
| rs13280055 | G/A | 1.08 (1.06-1.11) | 2.19*10^-11^ | 0.133 | rs13280055 | G/A | 1.12 (0.99-1.28) | 1.09*10^-01^ | - | rs13280055 | G/A | 1.12 (1.10-1.14) | 1.86*10^-07^ | - |
| rs1260326 | T/C | 1.08 (1.06-1.10) | 6.80*10^-18^ | 0.393 | rs1260326 | T/C | 1.20 (1.09-1.32) | 1.77*10^-04^ | - | rs1260326 | T/C | 1.08 (1.07-1.10) | 3.12*10^-08^ | - |
| rs6936023 | G/A | 0.94 (0.92-0.96) | 4.71*10^-08^ | 0.181 | rs6936023 | G/A | 1.01 (0.90-1.14) | 8.29*10^-01^ | - | - | - | - | - | - |
| rs192575995 | C/T | 1.26 (1.05-1.50) | 8.18*10^-09^ | 0.002 | rs192575995 | C/T | 2.40 (0.82-7.04) | 1.19*10^-01^ | - | - | - | - | - | - |
| rs2393775 | G/A | 1.09 (1.07-1.11) | 1.32*10^-23^ | 0.378 | rs2393775 | G/A | 1.08 (0.98-1.19) | 6.43*10^-02^ | - | rs2393775 | G/A | 1.08 (1.06-1.09) | 6.78*10^-08^ | - |
| rs17138478 | C/A | 1.09 (1.06-1.11) | 2.05*10^-11^ | 0.129 | rs17138478 | C/A | 1.04 (0.91-1.19) | 7.18*10^-01^ | - | rs17138478 | C/A | 1.10 (1.08-1.12) | 3.97*10^-07^ | - |
| rs1800961 | C/T | 1.28 (1.23-1.34) | 1.55*10^-29^ | 0.031 | rs1800961 | C/T | 1.33 (1.04-1.70) | 4.35*10^-02^ | - | rs1800961 | C/T | 1.46 (1.41-1.51) | 1.27*10^-30^ | - |
| rs753948177 | A/G | 1.67 (1.34-2.07) | 9.08*10^-09^ | 0.001 | rs753948177 | A/G | 1.44 (0.40-5.24) | 8.16*10^-01^ | - | - | - | - | - | - |
| rs12532734 | G/A,T | 0.92 (0.90-0.94) | 1.18*10^-16^ | 0.225 | - | - | - | - | - | rs17154498 | C/A | 0.91 (0.90-0.92) | 3.26*10^-10^ | 0.91 |
| rs542917327 | A/G | 1.33 (1.06-1.66) | 1.70*10^-08^ | 0.001 | rs542917327 | A/G | 4.80 (0.30-76.8) | 6.56*10^-01^ | - | - | - | - | - | - |
| rs34255979 | C/T | 1.13 (1.10-1.16) | 8.54*10^-20^ | 0.120 | rs34255979 | C/T | 1.02 (0.89-1.17) | 7.49*10^-01^ | - | rs34255979 | C/T | 1.11 (1.08-1.14) | 1.59*10^-04^ | - |
| rs367900721 | ATT/A | 1.05 (1.03-1.07) | 1.62*10^-08^ | 0.478 | - | - | - | - | - | - | - | - | - | - |
| rs2393969 | A/C | 1.07 (1.06-1.09) | 1.16*10^-17^ | 0.470 | rs2393969 | A/C | 1.07 (0.97-1.17) | 1.87*10^-01^ | - | rs2393969 | A/C | 1.06 (1.05-1.08) | 5.57*10^-06^ | - |
| rs749502751 | AC/A | 0.95 (0.93-0.97) | 4.67*10^-09^ | 0.367 | - | - | - | - | - | rs138430 | C/T | 0.97 (0.95-0.98) | 1.59*10^-02^ | 1 |
| rs56339318 | A/C | 0.92 (0.89-0.94) | 1.38*10^-09^ | 0.104 | rs56339318 | A/C | 0.87 (0.74-1.02) | 8.67*10^-02^ | - | rs56339318 | A/C | 0.92 (0.90-0.94) | 2.96*10^-04^ | - |
| rs528101230 | C/T | 1.39 (1.14-1.70) | 4.01*10^-08^ | 0.001 | - | - | - | - | - | - | - | - | - | - |
| rs3784924 | A/G | 1.06 (1.04-1.08) | 2.67*10^-11^ | 0.314 | rs3784924 | A/G | 1.10 (1.00-1.21) | 5.77*10^-02^ | - | rs3784924 | A/G | 1.07 (1.05-1.08) | 9.83*10^-06^ | - |
| rs764777280 | T/C | 1.33 (1.06-1.67) | 3.84*10^-08^ | 0.001 | rs764777280 | T/C | 0.33 (0.10-1.05) | 6.09*10^-02^ | 1.00 | - | - | - | - | - |
| rs2290846 | G/A | 1.13 (1.11-1.15) | 5.89*10^-38^ | 0.290 | rs2290846 | G/A | 1.08 (0.98-1.20) | 1.03*10^-01^ | - | rs2290846 | G/A | 1.13 (1.11-1.14) | 1.31*10^-13^ | - |
| rs2469991 | A/T | 0.94 (0.92-0.96) | 5.10*10^-10^ | 0.289 | rs2469991 | A/T | 0.98 (0.88-1.08) | 7.89*10^-01^ | - | rs2469991 | A/T | 0.97 (0.96-0.98) | 3.36*10^-02^ | - |
| rs3802548 | T/A | 1.12 (1.10-1.14) | 2.02*10^-29^ | 0.241 | - | - | - | - | - | rs3802548 | T/A | 1.10 (1.08-1.12) | 1.06*10^-10^ | - |
| rs10828250 | C/G | 1.06 (1.04-1.08) | 2.30*10^-10^ | 0.309 | rs10828250 | C/G | 1.01 (0.91-1.11) | 6.77*10^-01^ | - | rs10828250 | C/G | 1.03 (1.01-1.04) | 5.55*10^-02^ | - |
| rs7786376 | A/G | 1.05 (1.03-1.07) | 4.97*10^-08^ | 0.278 | rs2074768 | A/C | 0.94 (0.85-1.04) | 6.21*10^-01^ | 0.87 | rs7786376 | A/G | 1.03 (1.02-1.05) | 4.26*10^-02^ | - |
| rs183820313 | G/T | 1.38 (1.11-1.71) | 2.00*10^-08^ | 0.001 | - | - | - | - | - | - | - | - | - | - |
| rs190891554 | T/C | 1.38 (1.13-1.69) | 2.59*10^-08^ | 0.001 | - | - | - | - | - | - | - | - | - | - |
| rs28929474 | C/T | 1.36 (1.29-1.43) | 2.65*10^-30^ | 0.020 | rs28929474 | C/T | 1.29 (0.94-1.79) | 6.13*10^-02^ | - | rs28929474 | C/T | 1.48 (1.41-1.55) | 4.74*10^-16^ | - |
| rs60360195 | A/T | 1.06 (1.04-1.08) | 6.66*10^-09^ | 0.200 | rs60360195 | A/T | 1.02 (0.91-1.15) | 7.24*10^-01^ | - | rs60360195 | A/T | 1.04 (1.02-1.05) | 1.69*10^-02^ | - |
| rs4493564 | A/G | 1.05 (1.03-1.07) | 5.21*10^-09^ | 0.353 | rs4493564 | A/G | 0.98 (0.89-1.07) | 6.15*10^-01^ | - | rs4493564 | A/G | 1.05 (1.04-1.07) | 2.43*10^-04^ | - |
| rs144846334 | G/A | 1.35 (1.25-1.46) | 5.42*10^-17^ | 0.010 | rs144846334 | G/A | 1.47 (1.00-2.18) | 8.24*10^-02^ | - | rs144846334 | G/A | 1.05 (0.97-1.15) | 5.46*10^-01^ | - |
| rs438568 | A/G | 0.94 (0.93-0.96) | 4.85*10^-11^ | 0.394 | rs438568 | A/G | 0.91 (0.83-1.00) | 1.13*10^-01^ | - | rs438568 | A/G | 0.94 (0.92-0.95) | 5.47*10^-06^ | - |
| rs62129966 | C/A | 0.85 (0.83-0.87) | 1.06*10^-40^ | 0.164 | rs62129966 | C/A | 0.74 (0.65-0.85) | 5.35*10^-05^ | - | rs62129966 | C/A | 0.85 (0.84-0.87) | 7.68*10^-18^ | - |
| rs533560317 | A/G | 1.28 (1.05-1.57) | 1.09*10^-09^ | 0.001 | - | - | - | - | - | - | - | - | - | - |
| rs4681515 | A/G | 0.89 (0.88-0.91) | 2.84*10^-39^ | 0.437 | rs4681515 | A/G | 0.95 (0.87-1.04) | 3.36*10^-01^ | - | rs4681515 | A/G | 0.88 (0.87-0.90) | 1.36*10^-20^ | - |
| rs2292553 | G/A | 1.04 (1.02-1.06) | 1.22*10^-06^ | 0.437 | - | - | - | - | - | - | - | - | - | - |
| rs7599 | A/G | 0.95 (0.93-0.96) | 2.49*10^-11^ | 0.366 | rs7599 | A/G | 0.95 (0.86-1.05) | 2.60*10^-01^ | - | rs7599 | A/G | 0.94 (0.93-0.96) | 3.47*10^-05^ | - |
| rs11089985 | A/G | 1.05 (1.04-1.07) | 8.38*10^-10^ | 0.348 | rs11089985 | A/G | 1.05 (0.95-1.15) | 1.65*10^-01^ | - | rs11089985 | A/G | 1.06 (1.04-1.07) | 3.54*10^-05^ | - |
| rs686030 | C/A | 1.13 (1.11-1.16) | 1.96*10^-21^ | 0.142 | rs686030 | C/A | 1.06 (0.93-1.21) | 6.55*10^-01^ | - | rs686030 | C/A | 1.15 (1.13-1.18) | 3.49*10^-12^ | - |
| rs771297649 | CCTAAGTAT/C | 1.16 (1.09-1.22) | 3.82*10^-08^ | 0.021 | - | - | - | - | - | - | - | - | - | - |
| 2:234664586_ATC_A | ATC/A | 1.07 (1.05-1.09) | 1.71*10^-12^ | 0.312 | - | - | - | - | - | rs887829 | C/T | 1.09 (1.07-1.10) | 1.17*10^-09^ | 1 |
| Chr:Pos: Chromosome:Position, Maj/Min: Major allele/ Minor allele, MAF: Minor allele frequency, P Value: P value using allelic model, OR (95% CI): Odds ratio with 95% confidence interval, UKB: UK Biobank, GS-SFHS: Generation Scotland-Scottish Family Health Study.P values highlighted in bold reached the threshold of significance determined by Bonferroni correction. | | | | | | | | | | | | | | |

### Supplementary Figures

1. Supplementary Figure 1: Rectangular format Manhattan plot for the GWA Meta-analysis
2. Supplementary Figure 2: Truncated rectangular format Manhattan plot for the GWA Meta-analysis
3. Supplementary Figure 3: Rectangular format Manhattan plot for the UKB GWAS
4. Supplementary Figure 4: Truncated rectangular format Manhattan plot for the UKB GWAS
5. Supplementary Figure 5: Quantile-quantile plot
6. Supplementary Figure 6: Comparison of effect sizes between main GWAS and the “Procedure-only” subgroup analysis (participants with gallstones who did not require a surgical procedure were excluded from this analysis)
7. Supplementary Figure 7: Association with Serum Lipid Particles
8. Supplementary Figure 8: Association with Serum Liver Enzymes
9. Supplementary Figure 9: Association with Serum Bilirubin Fractions
10. Supplementary Figure 10: Association with Serum Blood Count
11. Supplementary Figure 11: Association with Serum Glucose and HbA1c
12. Supplementary Figure 12: Association with Serum C-Reactive Protein and Cystatin C
13. Supplementary Figure 13: Polygenic Risk Score Association with Phenotypic Traits (Part 1)
14. Supplementary Figure 14: Polygenic Risk Score Association with Phenotypic Traits (Part 2)
15. Supplementary Figure 15: Polygenic Risk Score Association with Phenotypic Traits (Part 3)

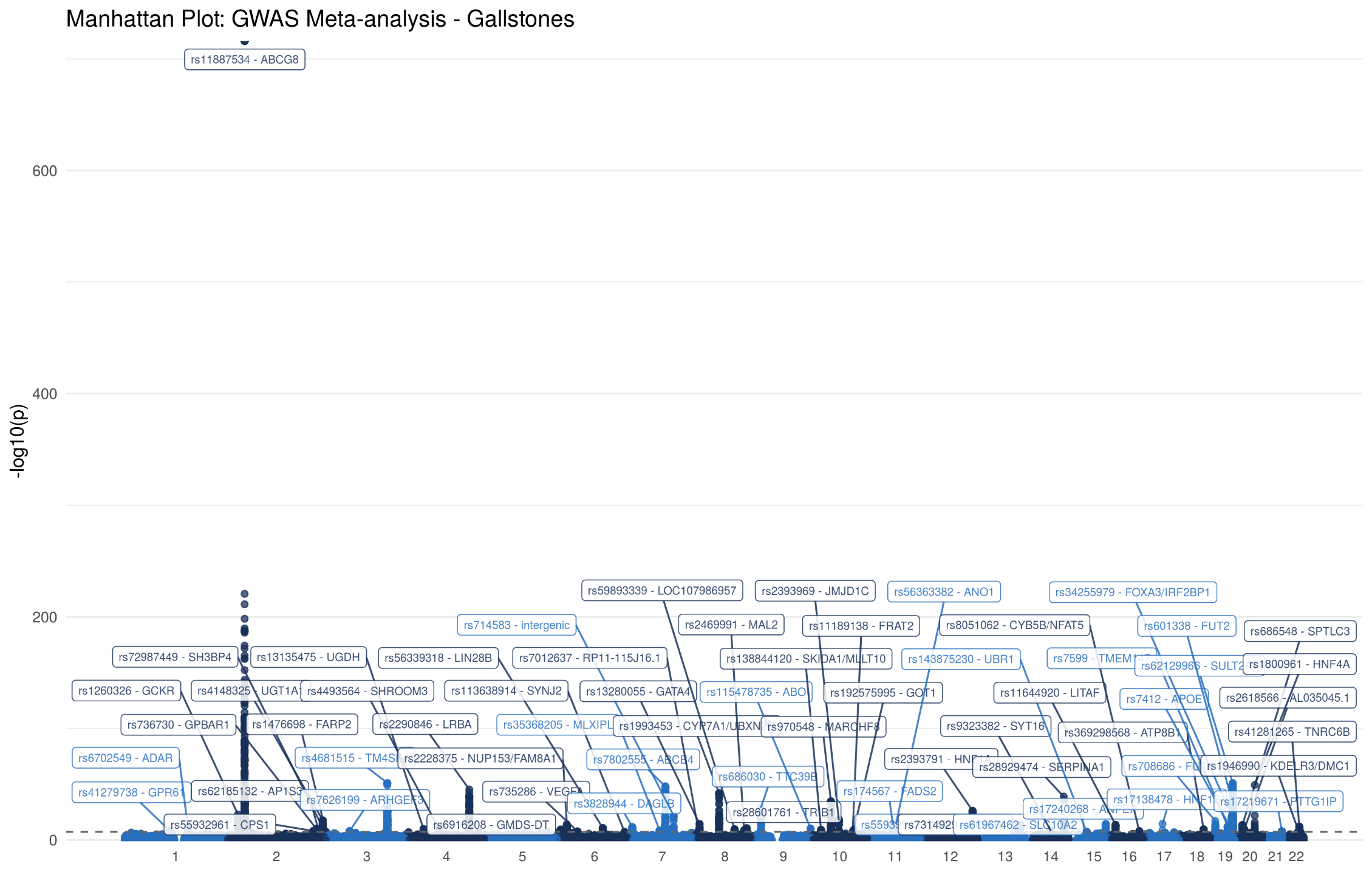

Supplementary Figure 1: Manhattan plot for the association with gallstone disease in the GWA Meta-analysis (43,639 cases and 506,798 controls). Each variant is plotted based on chromosome and position on the X-axis and -log10 P values on the Y-axis.

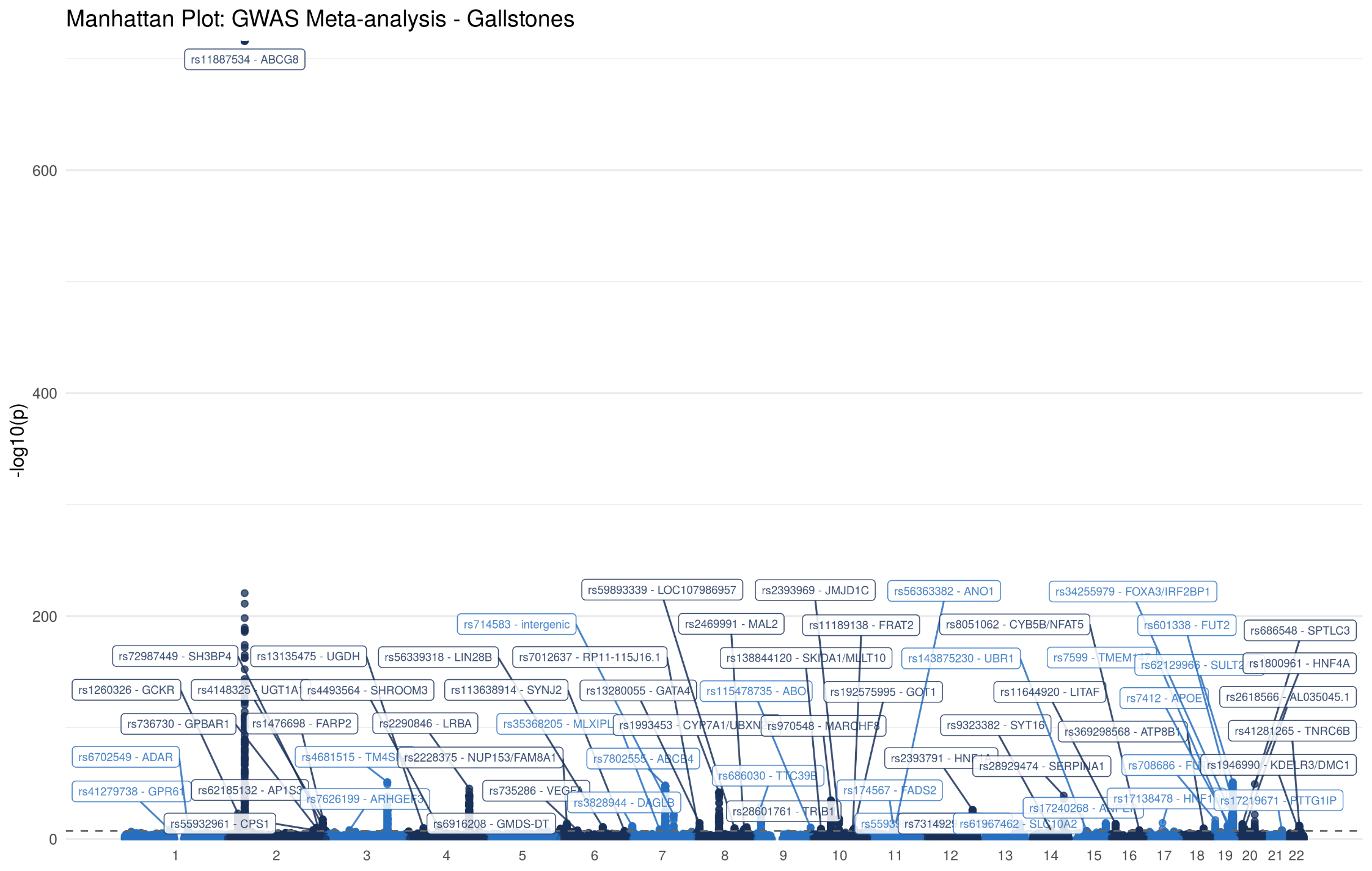
Supplementary Figure 2: Truncated Manhattan plot for the association with gallstone disease in the GWA Meta-analysis (43,639 cases and 506,798 controls). Each variant is plotted based on chromosome and position on the X-axis and -log10 P values on the Y-axis. Variants where $P<5*{10}^{-40}$ truncated.

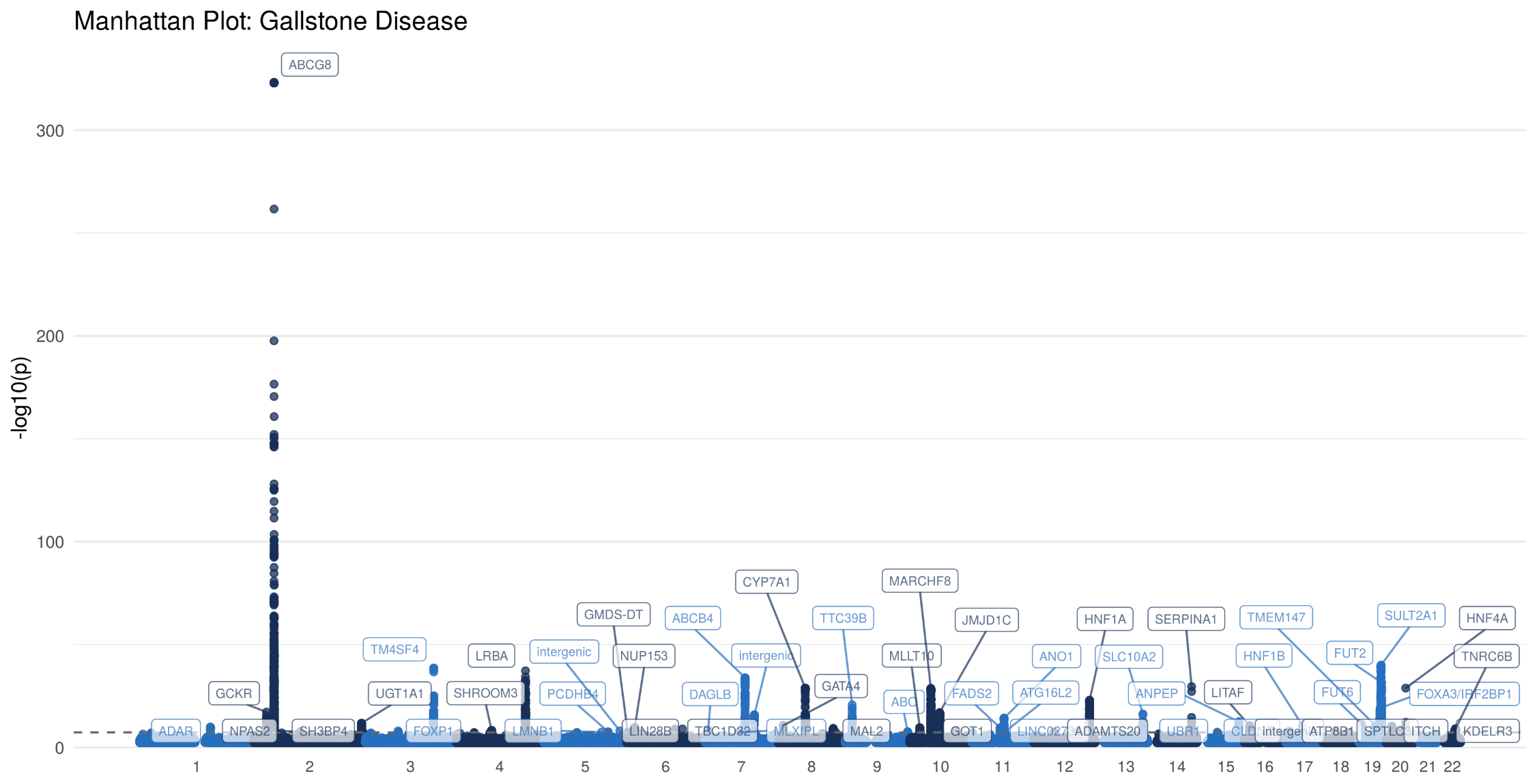

Supplementary Figure 3: Manhattan plot for the association with gallstone disease in the UK Biobank cohort (28,627 cases and 348,373 controls). Each variant is plotted based on chromosome and position on the X-axis and -log10 P values on the Y-axis.

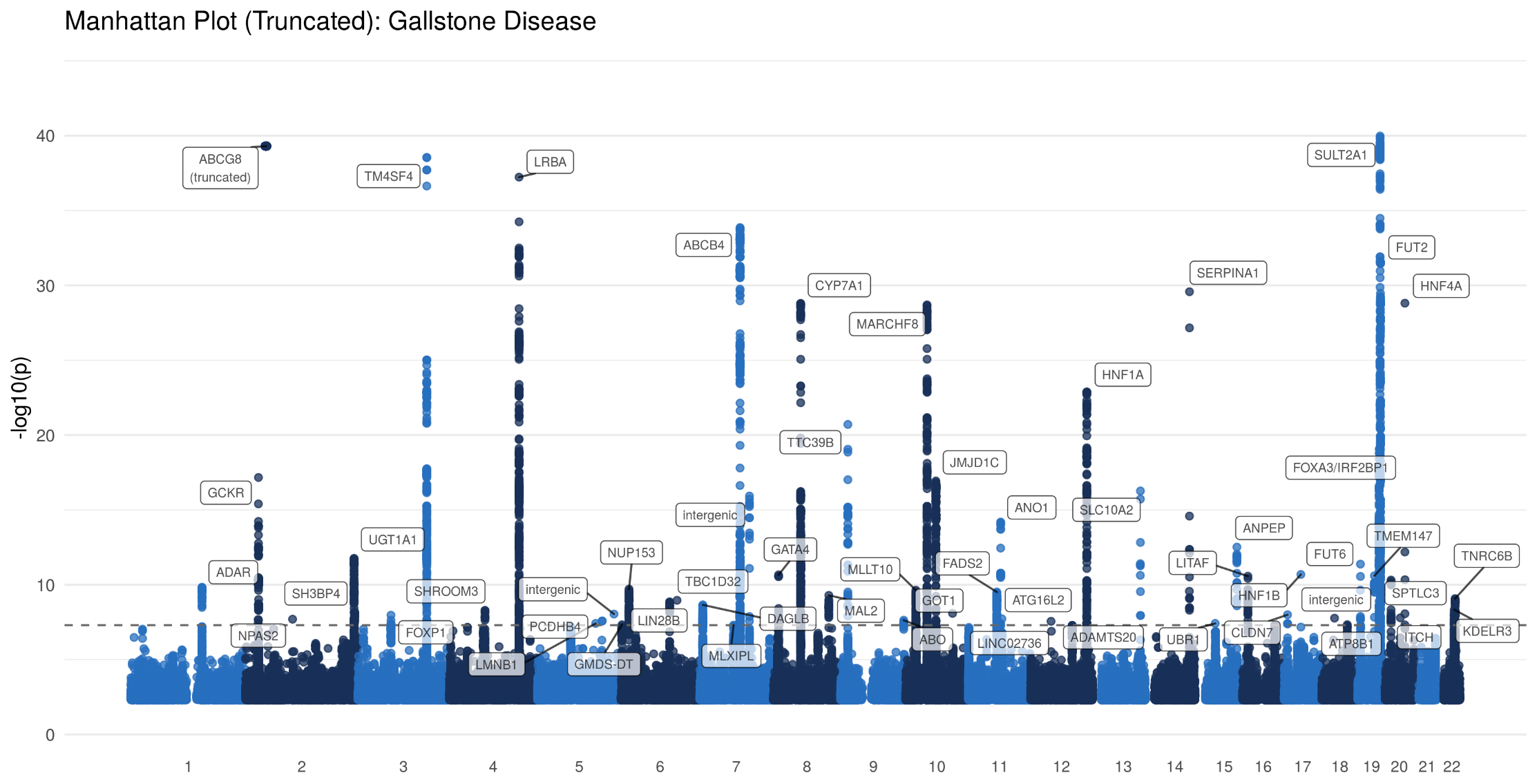

Supplementary Figure 4: Truncated Manhattan plot for the association with gallstone disease in the UK Biobank cohort (28,627 cases and 348,373 controls). Each variant is plotted based on chromosome and position on the X-axis and -log10 P values on the Y-axis. Variants where $P<5*{10}^{-40}$ truncated.

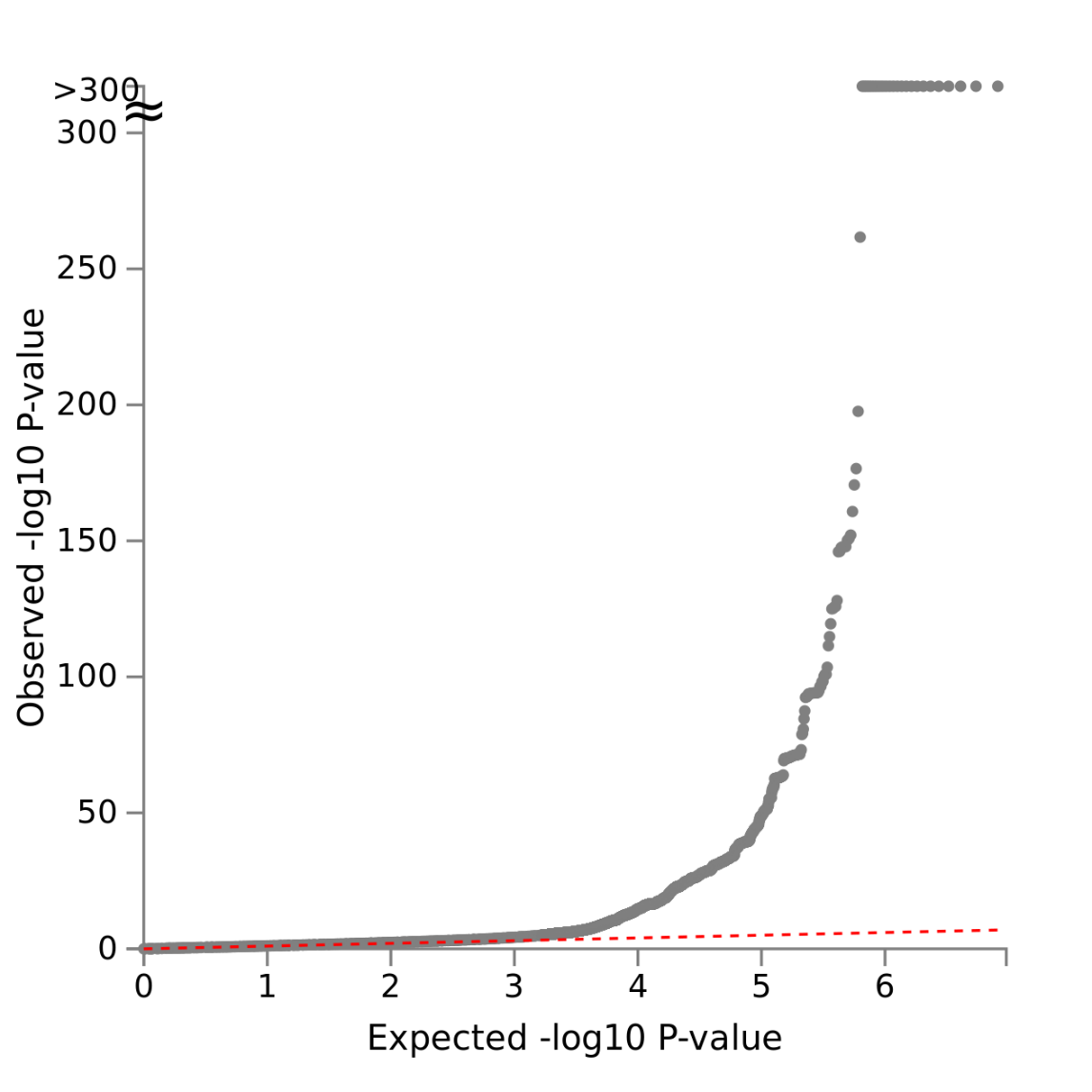

Supplementary Figure 5: QQ Plot showing the observed quantiles of gene effects on gallstones (y-axis) as a function of quantiles expected from a normal distribution (x-axis). Linkage disequilibrium regression score intercept 1.04.

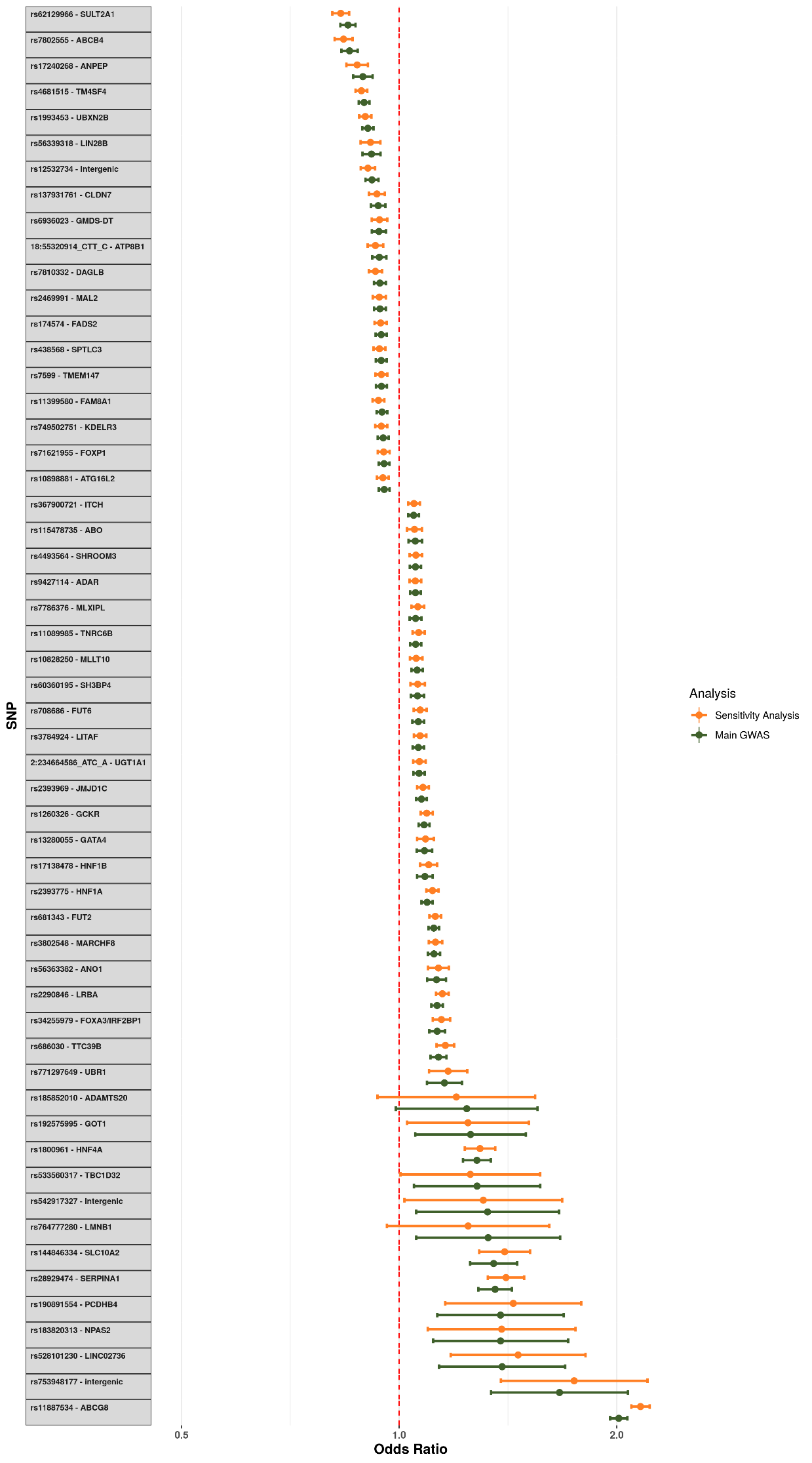

Supplementary Figure 6: Comparison of effect sizes at the lead SNP at each lead SNP between the main GWAS and the ‘Procedure-only’ sensitivity analysis. Each point represents the log odds for gallstones (main GWAS) or procedure for gallstones (sensitivity analysis) and the error bar represents the 95% confidence interval.

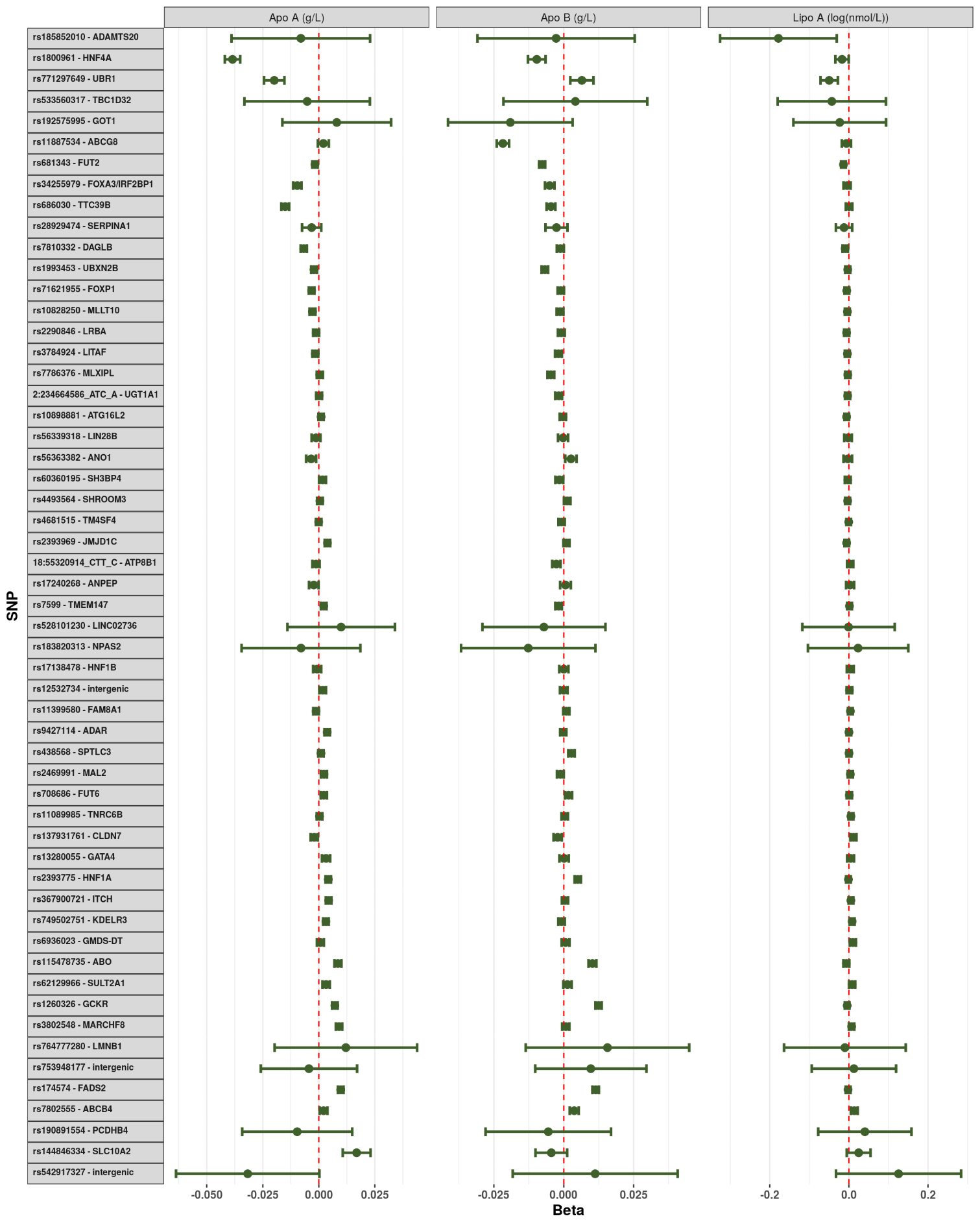

Supplementary Figure 7: Impact of each lithogenic allele on the measured serum lipid particles. Each point represents the beta-coefficient from an age- and sex-adjusted linear regression and the error bar represents the 95% confidence interval. Apo A - Apolipoprotein A; Apo B - Apolipoprotein B; Lipo A - Lipoprotein A. Lipoprotein A was log-transformed prior to analysis.

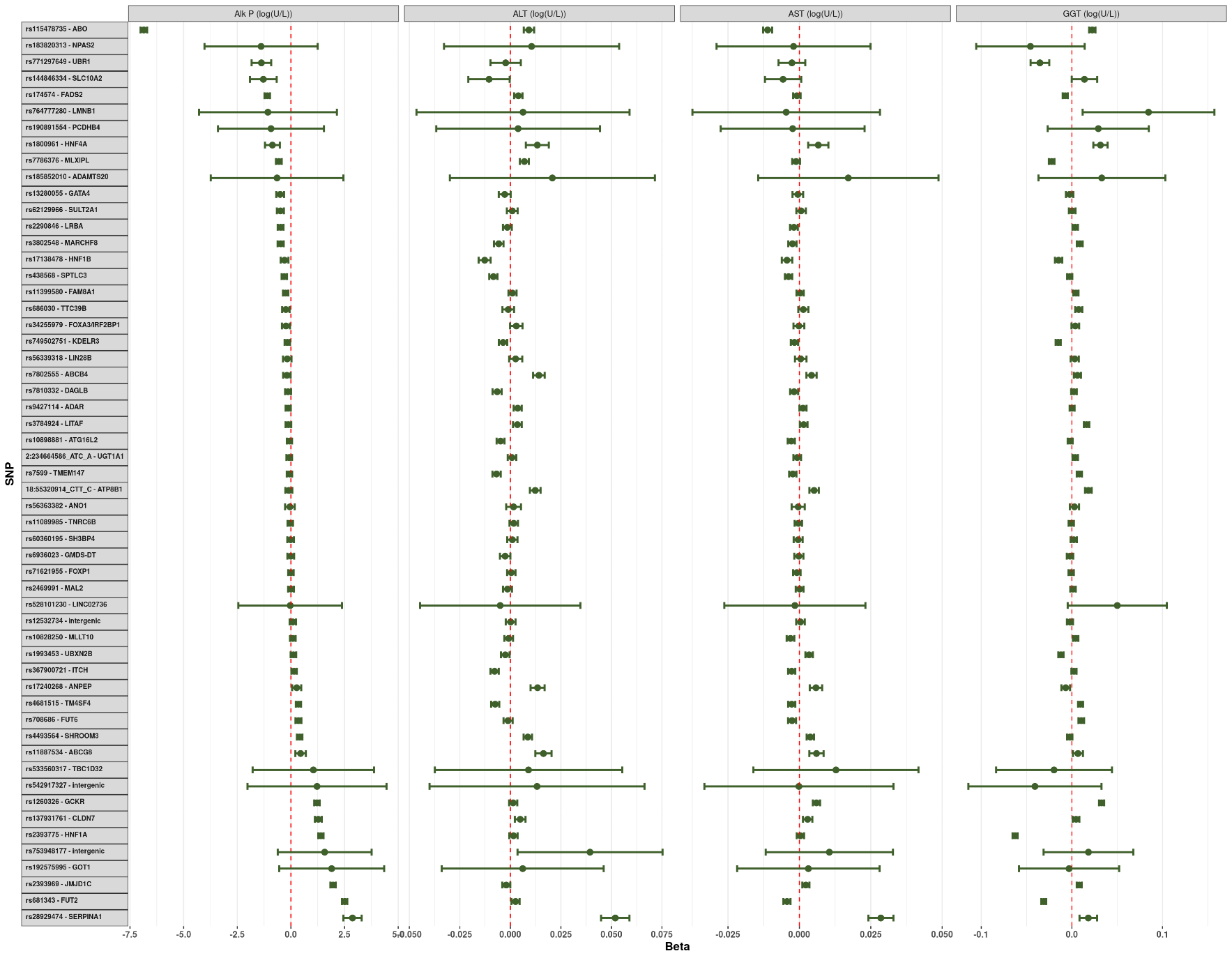

Supplementary Figure 8: Impact of each lithogenic allele on the measured serum liver enzymes. Each point represents the beta-coefficient from an age- and sex-adjusted linear regression and the error bar represents the 95% confidence interval. Alk P - Alkaline Phosphatase; ALT - Alanine Aminotransferase; AST - Aspartate Aminotransferase; GGT - Gamma Glutamyltransferase. Each of the serum biomarkers was log-transformed prior to analysis.

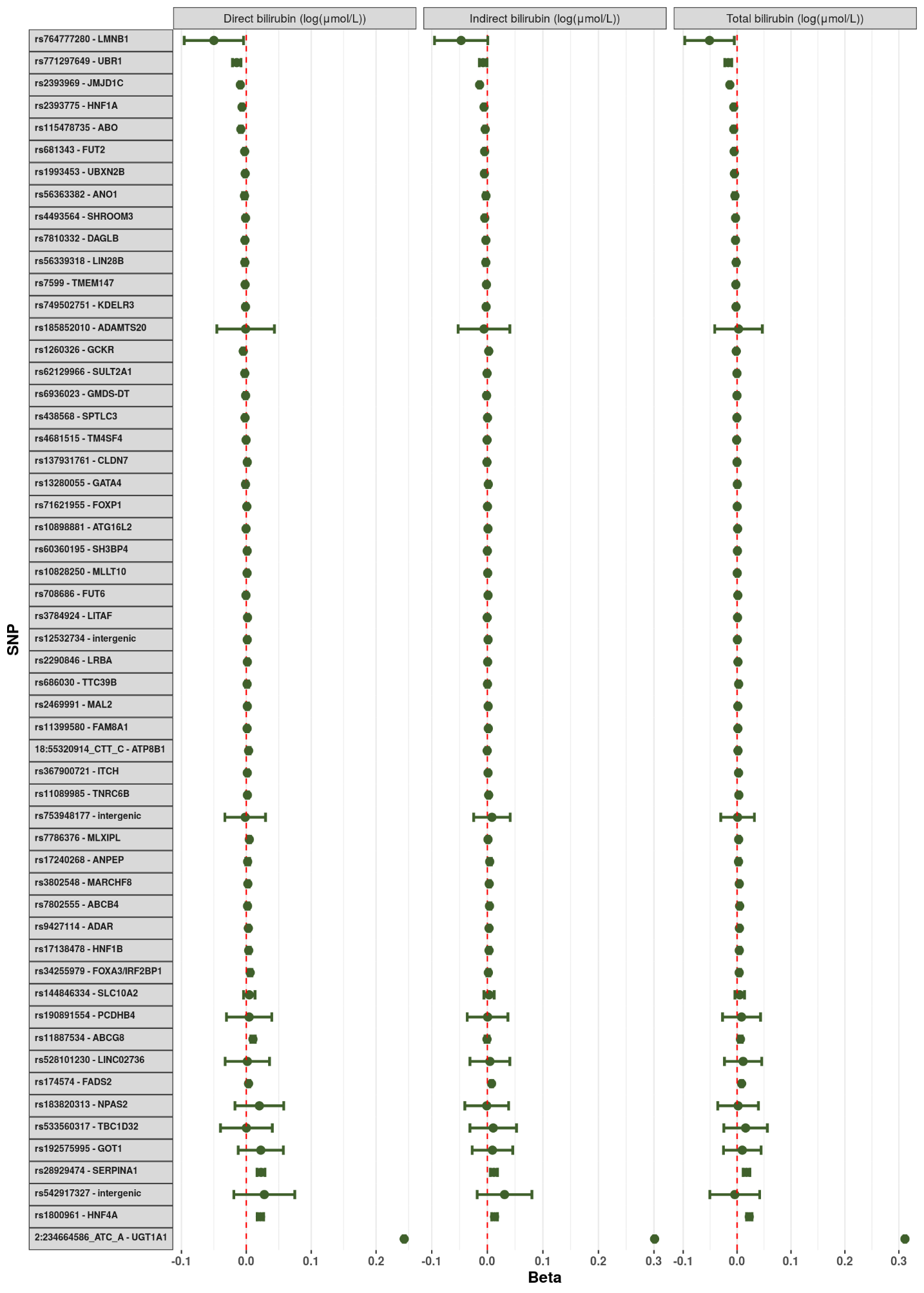
Supplementary Figure 9: Impact of each lithogenic allele on the measured serum bilirubin fractions. Each point represents the beta-coefficient from an age- and sex-adjusted linear regression and the error bar represents the 95% confidence interval. Each of the serum biomarkers was log-transformed prior to analysis.

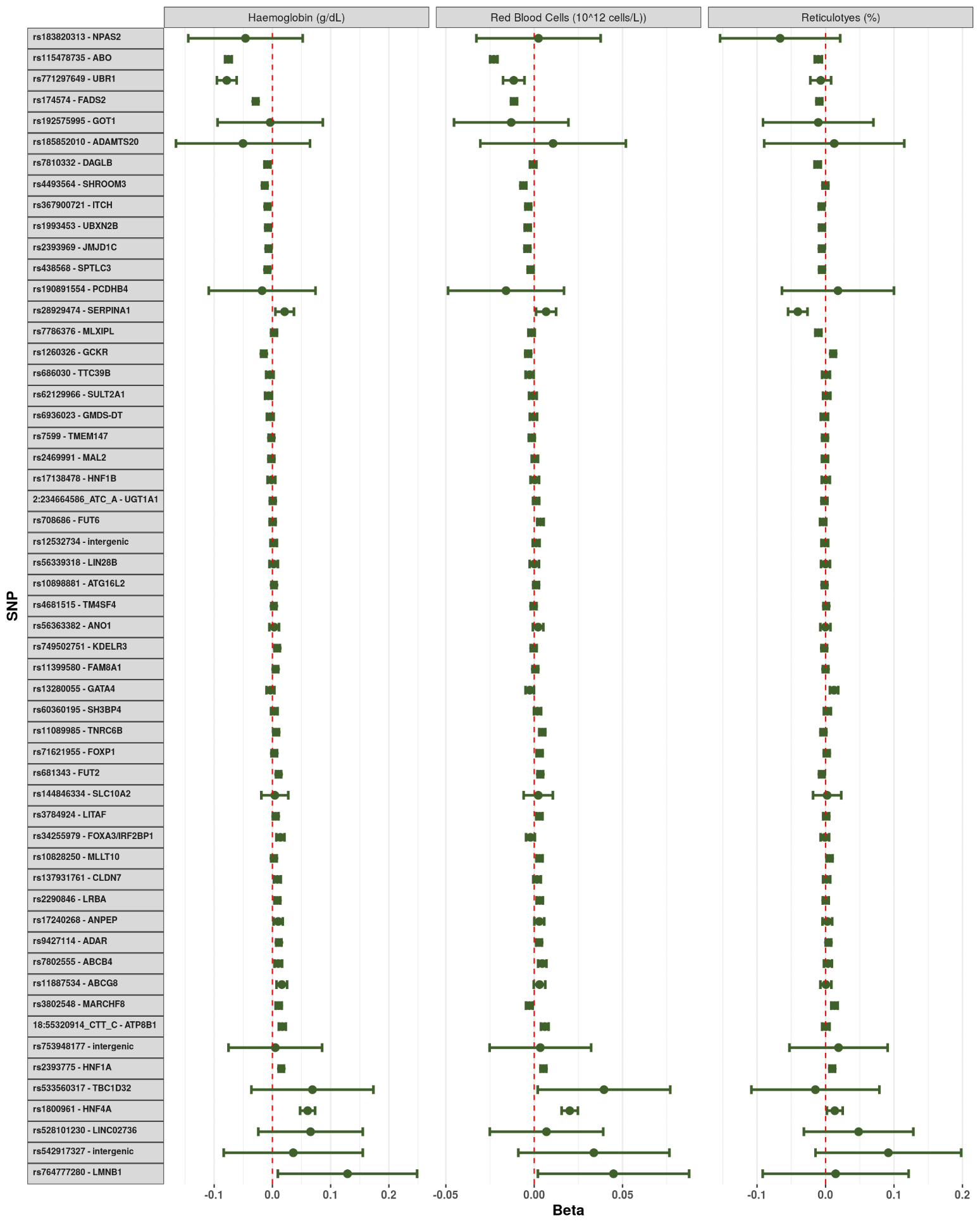

Supplementary Figure 10: Impact of each lithogenic allele on the measured serum blood count markers. Each point represents the beta-coefficient from an age- and sex-adjusted linear regression and the error bar represents the 95% confidence interval.

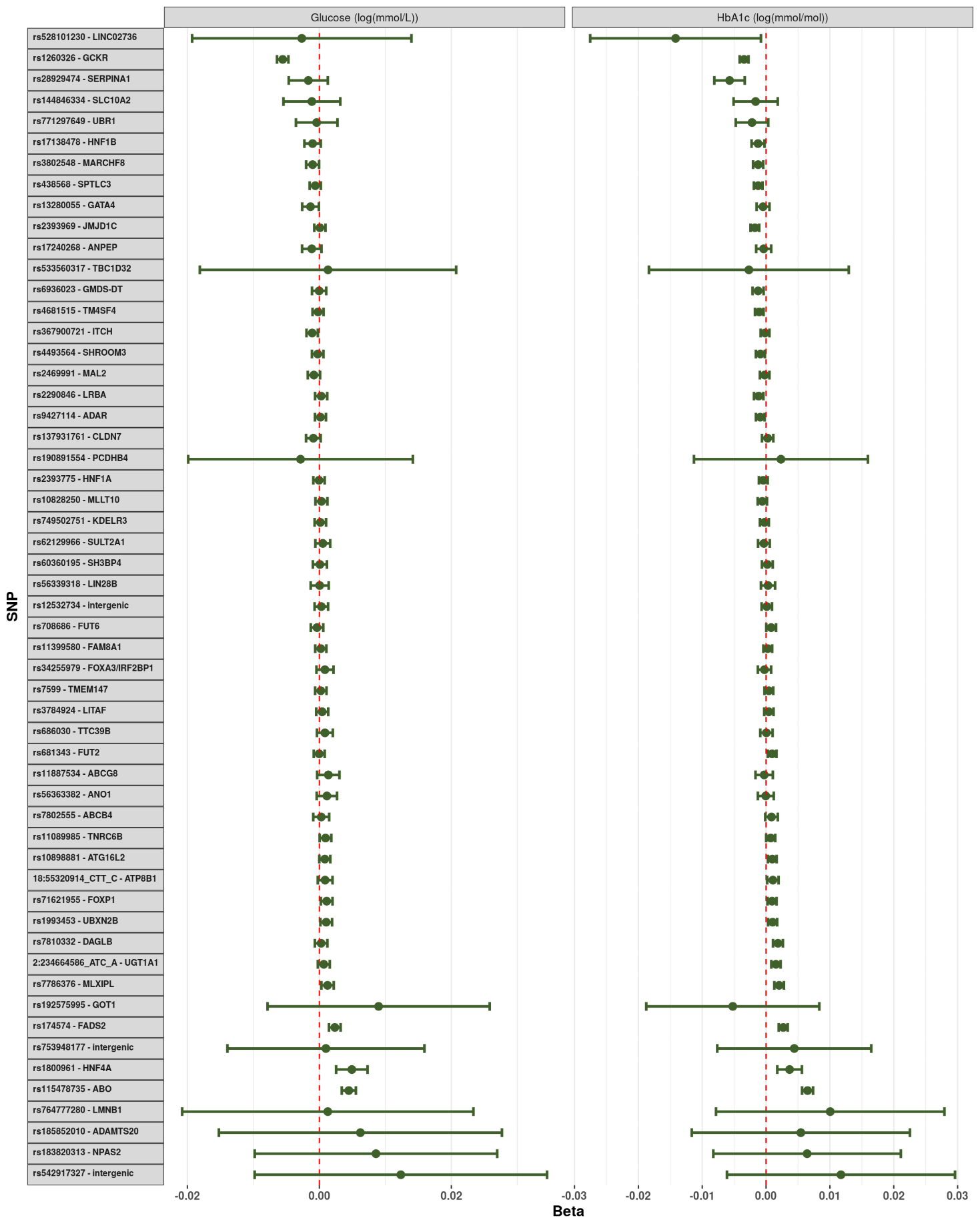

Supplementary Figure 11: Impact of each lithogenic allele on the measured serum glucose and glycated haemoglobin. Each point represents the beta-coefficient from an age- and sex-adjusted linear regression and the error bar represents the 95% confidence interval. HbA1c - glycated haemoglobin. Both glucose and HbA1c were log-transformed prior to analysis.

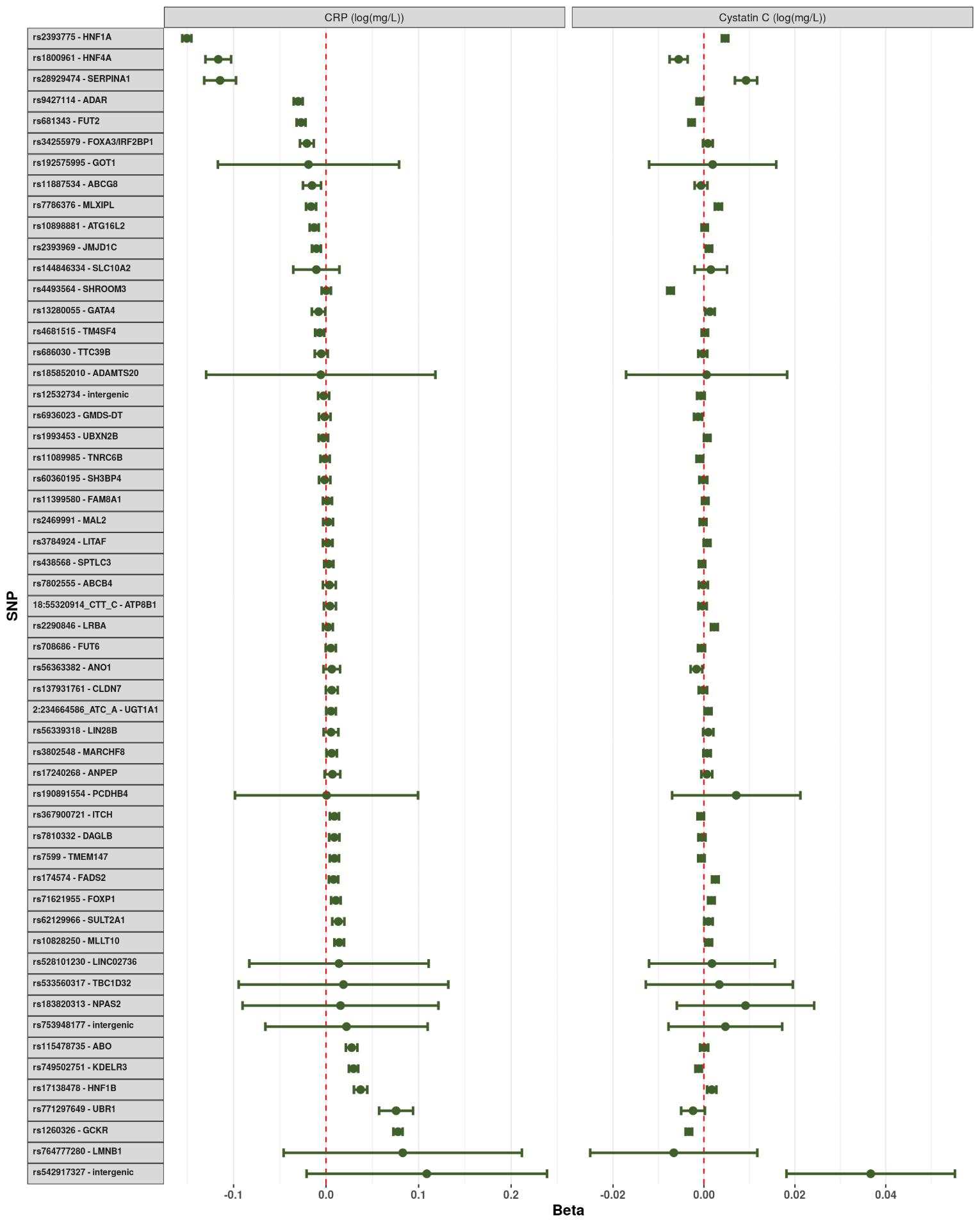

Supplementary Figure 12: Impact of each lithogenic allele on the measured serum C-reactive protein and cystatin C. Each point represents the beta-coefficient from an age- and sex-adjusted linear regression and the error bar represents the 95% confidence interval. CRP - C-reactive protein. Both biomarkers were log-transformed prior to analysis.

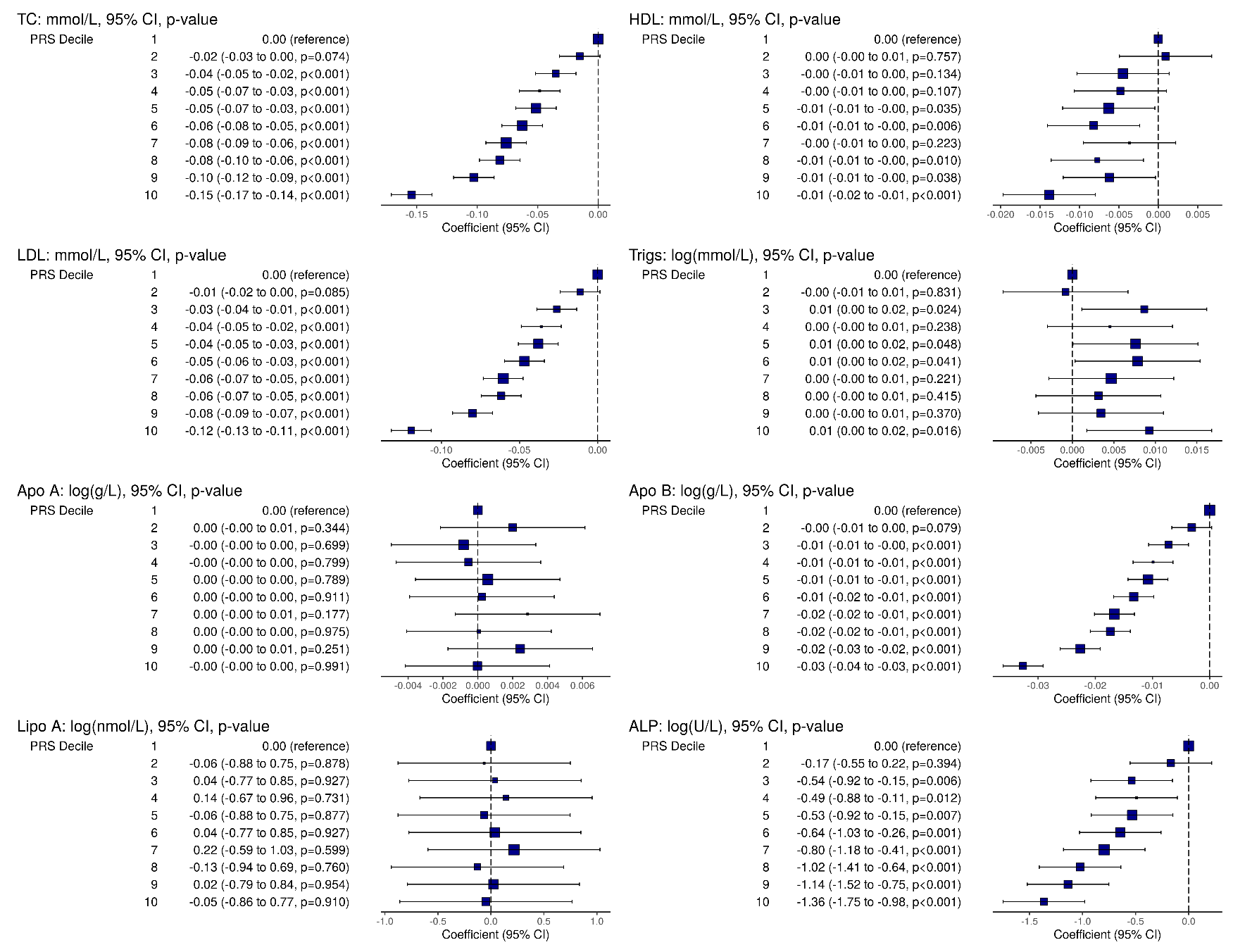

Supplementary Figure 13: Impact of each polygenic risk score on various phenotypic traits (Part 1). Each point represents the beta-coefficient from a simple linear regression and the error bar represents the 95% confidence interval. TC: total cholesterol, HDL: high-density lipoprotein, LDL: low-density lipoprotein, Trigs: triglycerdies, ApoA: apolipoprotein A, ApoB: apolipoprotein B, LipoA: lipoprotein A, ALP: alkaline phosphatase.

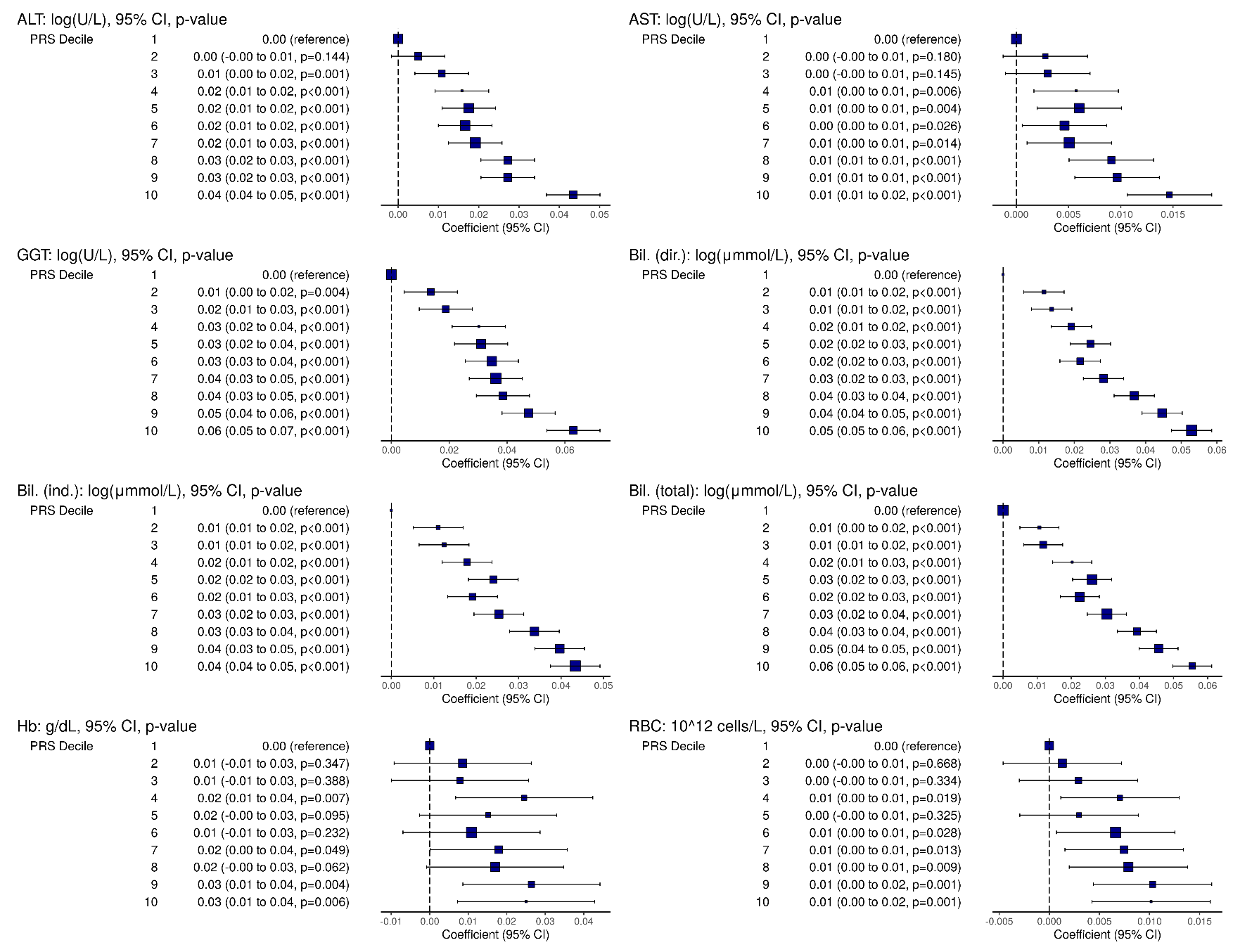

Supplementary Figure 14: Impact of each polygenic risk score on various phenotypic traits (Part 2). Each point represents the beta-coefficient from a simple linear regression and the error bar represents the 95% confidence interval. ALT: alanine aminotransferase, AST: aminotransferase, GGT: gamma glutamyltransferase, Bil. (dir.): direct bilirubin, Bil. (ind.): indirect bilirubin, Bil. (total): total bilirubin, Hb: haemoglobin, RBC: red blood cell count.

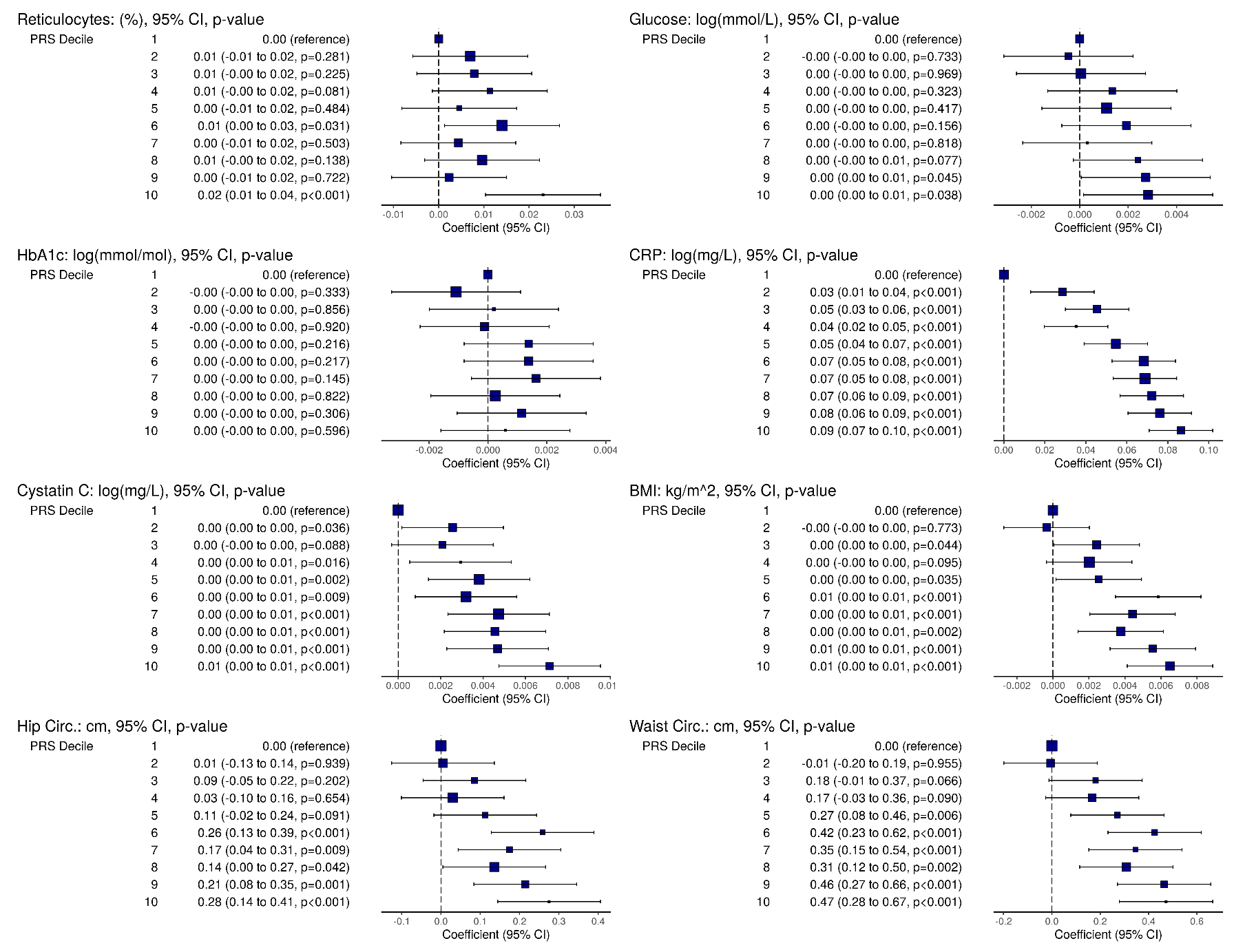

Supplementary Figure 15: Impact of each polygenic risk score on various phenotypic traits (Part 3). Each point represents the beta-coefficient from a simple linear regression and the error bar represents the 95% confidence interval. HbA1c: glycated haemoglobin, CRP: C-reactive protein, BMI: body mass index, Hip circ.: hip circumference, Waist circ.: waist circumference.
