## Supplementary Materials 2A for "Genome-wide Analysis Identifies Novel Gallstone-susceptibility Loci Including Genes Regulating Gastrointestinal Motility"

Supplementary Materials 2A, 2B, 4 and 5

Supplementary material including output for significant SNPs, FUMA results, sensitivity analyses, MAGMA results and other regression analyses has been made available through a separate interface as the files run to several thousands of lines and are too large to attach with the remaining Supplementary Materials.

The files are available for inspection at:

https://argoshare.is.ed.ac.uk/connect/#/apps/545/access
